# Optimal control of the spatial allocation of COVID-19 vaccines: Italy as a case study

**DOI:** 10.1101/2021.05.06.21256732

**Authors:** Joseph C. Lemaitre, Damiano Pasetto, Mario Zanon, Enrico Bertuzzo, Lorenzo Mari, Stefano Miccoli, Renato Casagrandi, Marino Gatto, Andrea Rinaldo

## Abstract

While SARS-CoV-2 vaccine distribution campaigns are underway across the world, communities face the challenge of a fair and effective distribution of limited supplies. We wonder whether suitable spatial allocation strategies might significantly improve a campaign’s efficacy in averting damaging outcomes. To that end, we address the problem of optimal control of COVID-19 vaccinations in a country-wide geographic and epidemiological context characterized by strong spatial heterogeneities in transmission rate and disease history. We seek the vaccine allocation strategies in space and time that minimize the number of infections in a prescribed time horizon. We examine scenarios of unfolding disease transmission across the 107 provinces of Italy, from January to April 2021, generated by a spatially explicit compartmental COVID-19 model tailored to the Italian geographic and epidemiological context. We propose a novel optimal control framework to derive optimal vaccination strategies given the epidemiological projections and constraints on vaccine supply and distribution logistic. Optimal schemes significantly outperform the explored alternative allocation strategies based on incidence, population distribution, or prevalence of susceptibles in each province. Our results suggest that the complex interplay between the mobility network and the spatial heterogeneities imply highly non-trivial prioritization of local vaccination campaigns. The extent of the overall improvements in the objectives grants further inquiry aimed at refining other possibly relevant factors so far neglected. Our work thus provides a proof-of-concept of the potential of optimal control for complex and heterogeneous epidemiological contexts at country, and possibly global, scales.

**Author summary:** The development of vaccines has sparked high hopes towards the control of SARS-CoV-2 transmission without resorting to extensive community-wide restrictions. A fundamental unanswered question concerns the best possible allocation of a limited vaccine stock in space and time given a specific goal. We address this through an optimal control framework based on a reliable spatially explicit COVID-19 epidemiological model, where vaccine distribution is optimized under supply and deployment capacity constraints. This tool provides strategies for optimal allocations in different scenarios, yielding important improvements over considered alternatives. By accounting for spatial heterogeneities and human mobility networks, the presented approach complements currently used allocation methods based on criteria such as age or risk.

## Introduction

Supply- or deployment-limited SARS-CoV-2 vaccines [1] pose the urgent question of a fair distribution of the available doses [2]. Current prioritization approaches typically target groups at higher risk of severe outcomes [3, 4], or their indirect protection by vaccinating those with higher disease transmission [3, 5, 6]. Our main hypothesis is that taking into account spatial heterogeneities in disease transmission when designing prioritization strategies significantly improves the effectiveness of vaccination campaigns. However, the distribution of doses inside each country is limited by the logistic capabilities of the healthcare network and the rate at which the vaccine stock is replenished. Decisions concerning the best allocation strategies are to be taken under these constraints. Moreover, both the complex coupling between regions due to human mobility and the spatial heterogeneities in disease history and control interventions make the discovery of such optimal allocation strategies an arduous task.

We propose an optimal control framework to explore COVID-19 vaccine distribution in space and time. We study the SARS-CoV-2 epidemic in Italy, where strong spatial effects arise from the geography of the disease, heterogeneous lockdown exit strategies, and post-lockdown control measures [7]. The optimal control framework is applied to a spatial model that has proved its reliability for Italy [8, 9], whose parameters are here sequentially updated through the assimilation of a year-long epidemiological record. This allows us to unravel the best possible vaccination strategy and probe the impact of vaccine allocations over the 107 Italian provinces.

The problem of vaccine allocation is of primary importance for public-health officials, epidemiologists, and economists [10, 11]. Roll-out strategies are conventionally based on the prioritization of individuals at risk, such as health workers and elderly people [12–15]. However, the heterogeneous ways in which different regions may be affected by each successive wave raise questions about spatial prioritization strategies. What is the best feasible spatial allocation, given supply and logistic constraints? Would that differ significantly from current non geographically-optimized plans? Should vaccines be distributed on the basis of demography or would it be better to prioritize areas currently subject to an outbreak? How relevant are the susceptibility profile and modelled future transmission in each region?

Epidemiological modeling has long been used to answer questions about the impact of vaccination campaigns, often by comparing outcomes under different scenarios [16, 17]. Optimization, i.e, the search for the best possible course of action that maximizes or minimizes an objective metric, has been carried out theoretically since the seventies [18–20]. Recent dramatic improvements of both algorithms [21] and computational power prompted applied studies using different methods to rigorously find optimal mitigation strategies [22–24]: most of the time trough iterative parameter search [25, 26], but also using genetic algorithms [27], greedy algorithms [28] or solving the Hamilton-Jacobi-Bellman equations [29, 30].

Interesting developments have recently arose during the ongoing SARS-CoV-2 pandemic [13, 31, 32]. The urgency of effective vaccination campaigns led to the development of modeling frameworks for the optimization of vaccine allocation, based on age or risk [3, 4, 12, 13], space [33], dose timing [34, 35], and the deployment of testing resources, using optimal control [36] or Bayesian experimental design [37], along with prioritization based on social contact networks [38].

To our knowledge, optimal spatial allocation of COVID-19 vaccines at a country scale has never been performed yet. This question is distinct from, and complementary to, risk-based prioritization. Spatial heterogeneities in disease transmission are complex, as seen during the initial outbreaks [8, 9, 39], supporting the significance of the posed problem towards an effective control of the epidemic. However, the connectivity network underlying spatial epidemiological models may generate complex large-scale control problems whose solution requires tailored formulations and efficient algorithms.

This work aims to find optimal strategies for this problem through modern optimization methods based on distributed direct multiple shooting, automatic-differentiation, and large-scale nonlinear programming [40–43]. This allows us to solve the large-scale optimization problems arising from epidemiological models, even when considering hundreds of spatial nodes.

## Materials and methods

The formulation of the optimal control problem has three main components: 1) an objective function to be minimized, here the number of new infections; 2) the spatial epidemiological model [8, 9] governing the transmission dynamics with the daily vaccination rates in each province as control variables; and 3) the set of constraints that the control must satisfy, in our case the limitations on vaccine administration rate in each province and the total vaccine stock in Italy.

### 1) Objective function

Optimizing calls for a metric, whose selection is critical in determining the optimal solution and its outcome. The choice of an objective function relates health, economy, and ethics. Possible candidates are the minimization of e.g. DALYs (the Disability-Adjusted Life Years), the number of deaths, disease exposure, and economic loss [44]. All these objectives are linked and may be combined together. As the model considered for this work does not have risk-classes, we optimize for the minimization of the incident infections in Italy from January 4, 2021 to April 4, 2021. Minimization of the deaths would yield the same results under the assumptions the model used.

### 2) Epidemiological model

Incidence and deaths are projected using the spatially distributed epidemiological model devised by Gatto et *al*. [8] and further improved by Bertuzzo et *al*. [9]. The model subdivides the Italian population into its 107 provinces represented as a network of connected nodes. Each province has local dynamics describing the number of individuals present in each of the model compartments: susceptible *S*, exposed *E*, pre-symptomatic *P* (incubating infectious), symptomatic infectious *I*, asymptomatic infectious *A*, hospitalized *H*, quarantined *Q*, recovered *R*, and dead *D*. A tenth compartment, vaccinated individuals *V*, is added to the original nine, as shown in Figure 1A.

**Fig 1.**
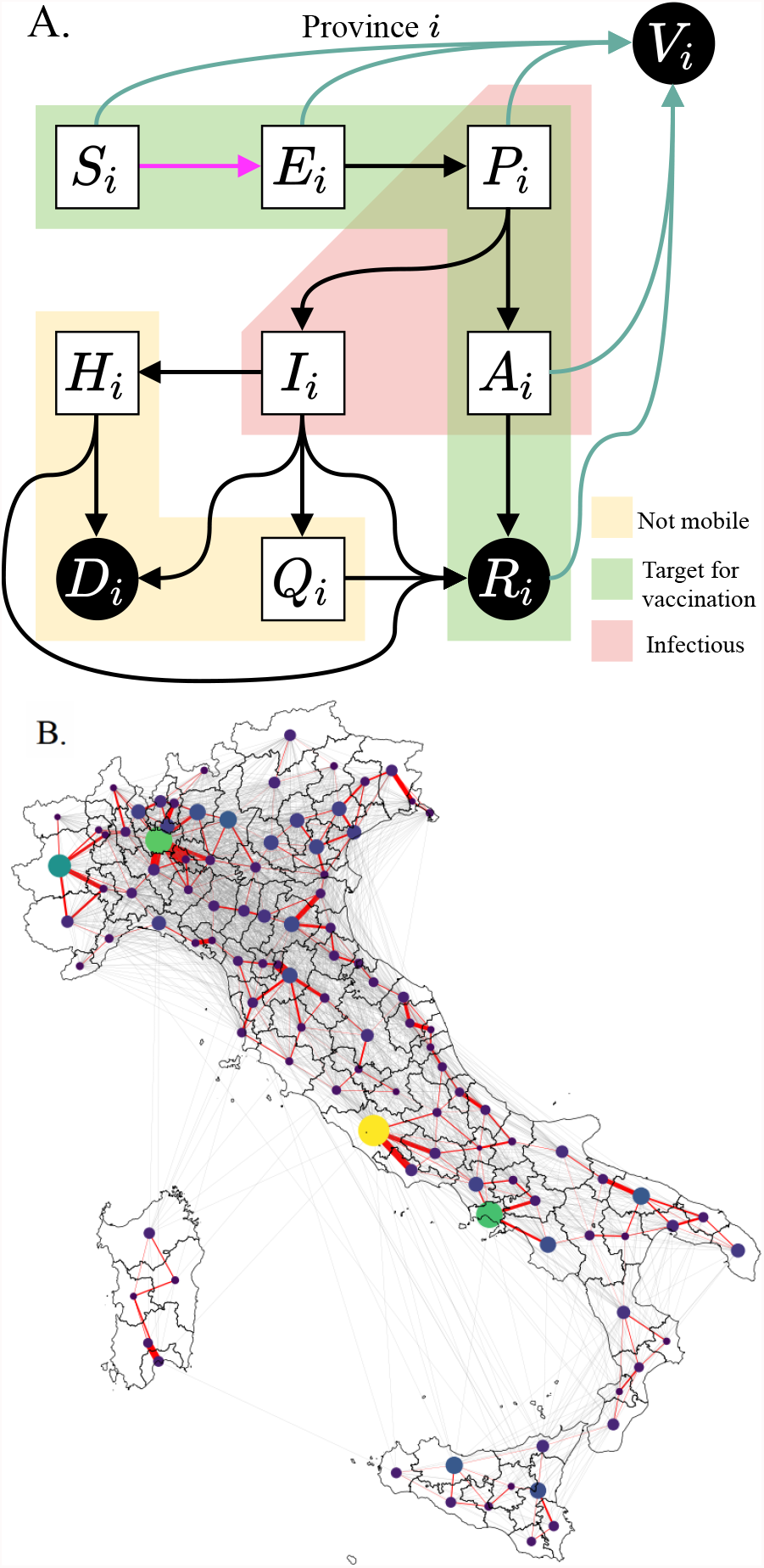
Model setup. (A) Diagram representing the compartments of the epidemiological model and the possible transitions in a single province. We control for the vaccination rate (teal arrows), aiming at minimizing incident infections (pink arrow). Individuals in compartments outside of the yellow block are able to move along the mobility network shown in (B), hence the force of infection in a province is coupled to the dynamics of other connected provinces. To reduce the problem to a tractable size, we only consider the most important connections (red edges) when optimizing, but we use the full network (red and grey edges) to assess our strategies. A discussion on the effect of this simplification is provided in SI. Nodes size and color display each province’s population, and edges width shows the straight of the coupling between each pair of province.

Except those in *H, Q, D* or *I* states, a fraction of individuals commutes between provinces along the mobility network, thus we introduce node-to-node disease transmission along the network shown in Figure 1B.

Compartments *P, A*, and *I* have different degrees of infectiousness and contribute to the force of infection (Equation SI (S3) and SI (S4)), which represents the rate at which susceptibles *S* become infected and, thus, enter the exposed compartment *E*. The force of infection in each province has a local and a mobile component. The local component describes transmission among the individuals that do not leave the node. The mobile component considers that local susceptibles may enter in contact with infected individuals that are traveling, and oppositely, susceptible commuters may become infected through contact with local infected. Connected provinces contribute to this process depending on the strength of the mobility fluxes from and to the node of interest. These mobility fluxes change in time due to the governmental policies introduced to reduce transmission among regions (more details about the data used to construct the mobility network and its use in the model are presented in the SI and in [8]).

The epidemiological model, previously calibrated during the first wave of COVID-19 in Italy [8, 9], is updated up to January 4, 2021 using an iterative particle filtering, which infers the regional transmission on a moving temporal window of two weeks. This data assimilation scheme allows us to capture the second wave of infections that hit Italy in the Fall of 2020, a necessary requirement to generate model projections that take into account the whole epidemic history, as shown in Figure 2. In our approach, model projections are described by an ensemble of a hundred trajectories associated with different parameters, whose distributions quantify the model uncertainty. We consider two projection scenarios characterized by two possible rates of epidemic transmission, see Figure 2. The “Optimistic” scenario assumes a constant lowering of transmission from January 4, 2021 to April 4, 2021; the “Pessimistic” scenario considers a gradual increase in transmission until mid February 2021, which results in a third wave.

**Fig 2.**
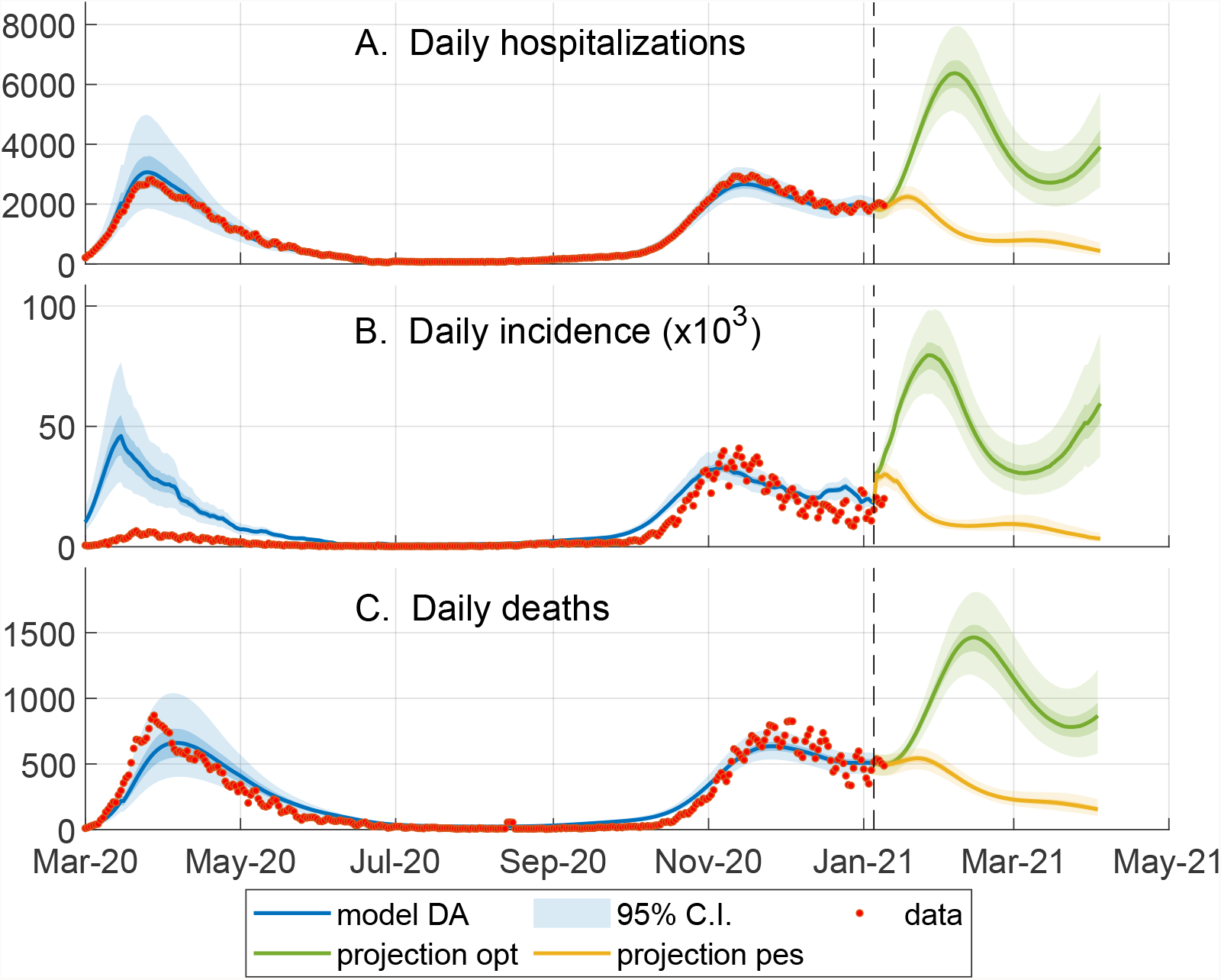
Data assimilation and scenarios for optimization. Comparison between the model outputs (95% confidence interval (CI) of the ensemble, blue shaded area) and the corresponding epidemiological data (red circles, obtained from the bulletins of the Dipartimento della Protezione Civile, https://github.com/pcm-dpc/COVID-19) from March 2020 to January 2021. The orange and green shaded area respectively show the ensemble dynamics (95% CI) of what we called pessimistic and optimistic transmission scenarios from January to April. The optimal vaccination strategy in the optimistic (or pessimistic) scenario is computed with respect to the the continuous green (or orange) line, representing the model trajectory obtained using the median of each ensemble parameter. (A) The data on the daily hospitalizations is estimated as described in [9]; this data at the regional level is assimilated on a moving window of 14 days to update the model parameters describing the local transmission rates (see SI). (B) Daily number of newly exposed individuals versus the reported positive cases. Note that the large discrepancy between model and data during the first wave is due to the low testing capacity at the beginning of the epidemic (C) Daily number of deaths.

The control variable is the vaccination rate in each province. We assume one-dose vaccines with instantaneous 100% efficacy while in reality the vaccine efficacy and immunity duration depends on the vaccine type. As we focus on spatial patterns and differences among vaccination strategies for a given supply, this assumption does not affects the conclusion of our work. Finally, we impose that vaccine protection persists during the three months of projection considered.

For each scenario, the optimal control problem is solved for one reference model trajectory, whose parameters and state on January 4, 2021 are obtained as the median values of the 100 model realizations. In this way, the reference trajectory approximately represents the ensemble median in each province. Then, we assess the effectiveness of the optimal allocation on the full ensemble of trajectories.

### 3) Constraints

We define two types of constraints: supply constraints, which determine the weekly delivery to the national stockpile; and logistic constraints, which limits the maximum rate of local vaccine distribution in each province.

The supply constraints ensure that the model does not distribute more vaccine than what is actually available in stock. We assume that the national supply of vaccine doses is empty on January 4, 2021 and is replenished every Monday. We consider four scenarios with weekly deliveries of 125’00, 250’000, 479’700 (realistic, baseline value) and 1M vaccine doses (additional scenarios with 1.5M and 2M doses delivered are shown in SI).

From the national stockpile, doses may be allocated to any of the 107 Italian provinces, but the logistic constraints limit the rate at which it is possible to distribute the vaccines in each province. We assume that the maximum number of individuals who can be vaccinated in a province per day is proportional to the province’s population, such that the national maximum distribution capacity equals 500,000 doses per day, i.e., 3.5M per week if every province vaccinates at its maximum rate (which in retrospect is close to Italy’s vaccination rate as of May 1st).

The objective, the model, and the constraints may be tailored to specific applications within the proposed framework.

Using state-of-the-art linear algebra solvers and automatic differentiation, we solve each scenario (optimistic and pessimistic, with different weekly stockpile deliveries) for the optimal vaccination allocation.

## Optimal control problem formulation

We provide a brief methodological description of the optimal control framework. The full equations are derived in the SI, along with implementation details and the source code.

We denote *n* the number of spatial nodes (*n* = 107 provinces in Italy) and *m* the number of epidemic states in our model (*m* = 9 states). We denote as 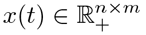 the state of the system, i.e., *x*(*t*) is a vector containing the epidemic variables *S*_*i*_(*t*), *E*_*i*_(*t*), *P*_*i*_(*t*), *I*_*i*_(*t*), *A*_*i*_(*t*), *Q*_*i*_(*t*), *H*_*i*_(*t*), *R*_*i*_(*t*), *V*_*i*_(*t*) for every province *i* = 1, …, *n*. We define 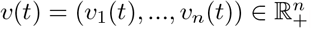, representing the rate of vaccine rollout for every node *i* at time *t*, as our control variable. The epidemiological model can be described by the following system of ordinary differential equations coupling disease transmission among all provinces:

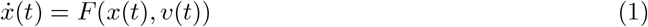

The national incidence, i.e., the sum of new infections in all provinces at time *t*, is selected as the cost function, *L*(*x*(*t*), *v*(*t*)). Given our system with states *x* subject to the dynamics (1) and controls *v*, the optimal control problem is formalized as:

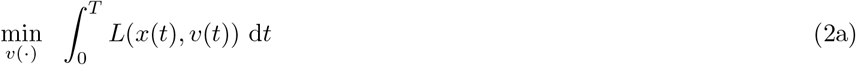

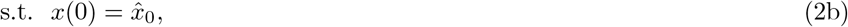

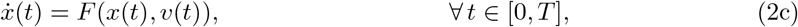

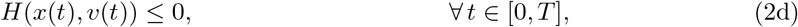

where we aim at minimizing the cost function over the control horizon *T*, while enforcing the modeled SARS-CoV-2 transmission dynamics (Equations (2b) and (2c)). Moreover, the constraints imposed by vaccine availability and the maximum vaccination rate are lumped in function *H* that expands to

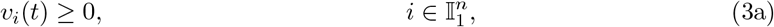

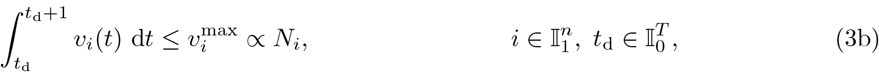

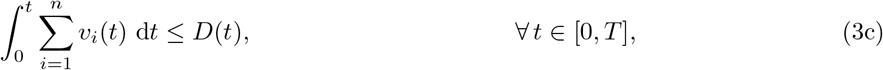

where time is measured in days, and 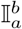 is the set of all integers *a* ≤ *k* ≤ *b*. Equation (3a) enforces that one can only distribute a non-negative amount of vaccine doses. Equation (3b) states the logistic constraints, which limit to 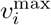 the amount of individuals that can be vaccinated each day in each node; here *t*_d_ is the time at which each day starts. We impose that the daily vaccination capacity of each province is proportional to its population size *N*_*i*_, assuming a fair distribution of the sanitary infrastructure among provinces, as shown in SI (Figure S2). The constraint on the national stockpile is materialized by Equation (3c), which ensures that the total vaccine allocation across all nodes does not exceed the stockpile *D*(*t*). The stockpile is replenished every Monday by the delivery of new vaccines, hence *D*(*t*) is a staircase function. For an overview of the possible solution approaches for optimal control problems we refer the interested reader to [45, 46]. In particular, in this work, we use a variant of direct multiple shooting [40] tailored to distributed systems [41]. We solve the optimal control problem in Equation (2) by a direct method, also called *first discretize, then optimize*, which transforms the control problem into a nonlinear programming problem. We split our time window [0, *T*] into *N* intervals [*t*_*k*_, *t*_*k*+1_], and we denote as *x*_*k*_ = *x*(*t*_*k*_) the states at time *t*_*k*_, and as *v*_*k*_ the controls in interval [*t*_*k*_, *t*_*k*+1_]. The continuous-time dynamics *F* in Equation (1) are transformed by numerical integration into the discrete-time operator *f* by numerical integration. This discretization requires some care, and details are provided in the SI. We thus obtain the following nonlinear programming problem:

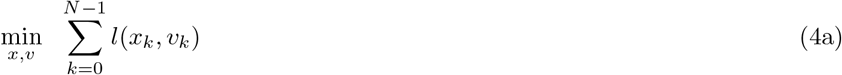

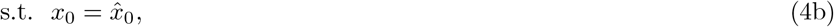

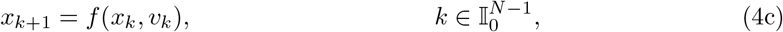

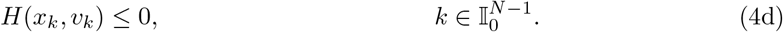

Nonlinear programming problems may be solved by readily available solvers using the primal-dual interior point method. The main difficulty in solving the proposed nonlinear programming problem (4) is the large dimension of the system and the non-linearity of the model. In order to bring the problem to a tractable form, we introduce three simplifications: (a) vaccines are administered instantaneously at the beginning of each day, rather than with a constant rate over the whole day; (b) the component of the force of infection taking into consideration the mobility of individuals across provinces is evaluated at the beginning of each day and remains constant through the day; and, (c) the mobility network is simplified, by keeping only the most important connections (see Figure 1), thus increasing the sparsity of the underlying spatial connectivity matrix. These simplifications deliver a significant computational advantage, and we verified that the impact on the model accuracy is limited. Note that, even if the optimal strategy is computed using the simplified model, its impact in terms of averted infections (shown in **Results**) is evaluated using the full epidemiological model without any of these simplifications. A more detailed discussion on this subject is provided in the SI.

The nonlinear programming problem arising from the simplified epidemiological model is non-convex, and involves approximately 10^5^ variables and 10^5^ constraints. We formulate the problem using CasADi [42] and solve it using Ipopt [43] with sparsity-exploiting linear algebra solvers. In practice, solving the optimal control problem takes between two to four days on a 36-cores 2.3 GHz CPU.

## Results

We obtain the optimal vaccination strategies for a set of eight scenarios drawn from the spatial model from January 4, 2021 to April 4, 2021. These scenarios are a combination of two projection scenarios (pessimistic vs optimistic) and four assumptions on the weekly stockpile delivery (125’000, 250’000, 479’700 or 1M doses delivered per week). In each scenario, the optimal solution is a spatially explicit vaccine roll-out policy, i.e. an indication of the number of vaccine doses to be deployed in each province each day.

### Comparison of vaccination strategies

Spatial prioritization based on epidemiological criteria, such as past [16] or future [17] incidence, has been thoroughly used in real campaigns and prospective studies.

In order to measure the potential impact of the optimal allocation strategy, we compare it against 12 alternative approaches which distribute the available weekly vaccine doses among provinces.

These strategies uses an indicator variable to rank provinces, either i) their population; ii) the number of susceptible (per inhabitant or absolute) individuals at the beginning of the projection; iii) the future incidence (per inhabitant or absolute) as projected by the epidemiological model or iv) constant, equal for all province. The incidence indicator ranking is updated to reflect the change of each past decision. Then these strategies either focus on the provinces where the indicator is the highest or allocate to all provinces proportionally to the indicator. Additionally, a greedy strategy from the literature is presented [28]. In the main text, we present the results of optimal strategy, the second best strategy overall (with indicator incidence per inhabitant and focused allocation) and proportional strategy for incidence, susceptibility and population; the other analysis are left in the SI.

For each of the eight scenarios considered, we compute the number of averted infections with respect to a zero-vaccination baseline, and the number of averted infections per vaccination dose (see Table 1). In the optimistic transmission scenario, characterized by a recess of the epidemic, the vaccination campaign has a lower impact on the averted infections per dose as only a small percentage of the vaccinated individuals would have been at risk of transmission. As expected the impact of the vaccination campaign is more evident in the pessimistic scenario where the optimal strategy averts up to 2.54 million infections given a weekly stockpile deliveries of one million doses. By virtue of the law of diminishing returns, the number of averted infections per dose decreases (from 0.413 to 0.196) when increasing the weekly stockpile.

**Table 1.**
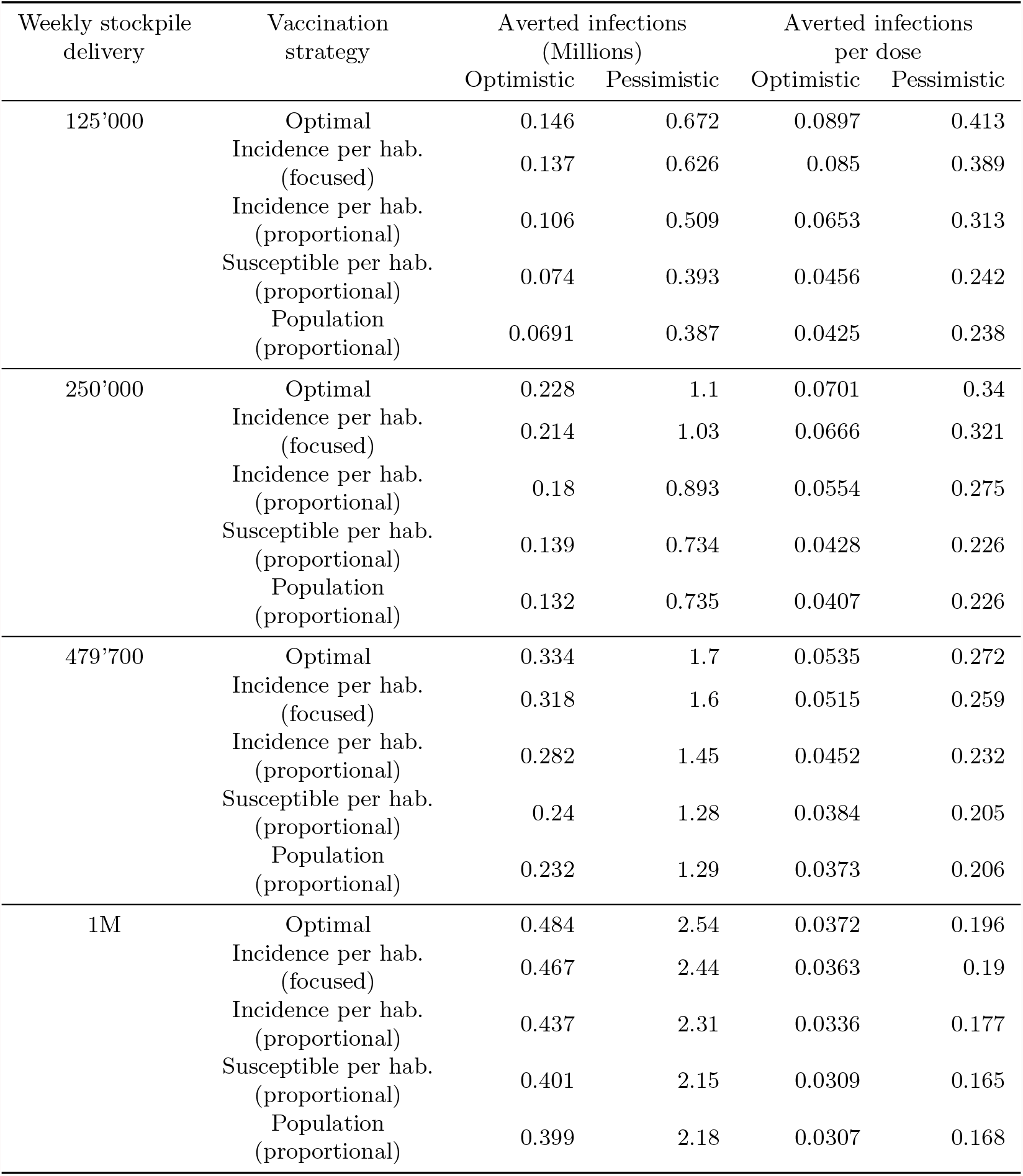
Averted infections. Absolute number of averted infections and averted infections per dose during the first three months of 2021 as evaluated for the reference trajectory (see Figure 2). The first column represents the considered scenarios of weekly stockpile replenishment, i.e. the number of doses delivered to Italy every week, ranging from 125’000 to one million.

The optimal solution always outperforms all the explored alternative strategies in terms of the number of averted infections and in terms of averted infections per dose allocated (see Tables 1, 1, and 2). The alternative strategy focusing on the provinces with the largest incidence has results close to the optimal strategy, with a difference of less than 10% in each scenario. Instead, other strategies are significantly less effective. The improvement between optimal and incidence-based (proportional) allocation is significant, ranging from 9.0% (pessimistic, 1M doses/week) to 27.4% (optimistic, 125’000 doses/week). In Figure 3, the black diamonds represent the percentage of averted infections obtained using each strategy for the reference trajectory, with respect to the averted infections resulting from the optimal strategy. We observe that, in both the optimistic and pessimistic scenarios, the optimal strategy has the largest relative benefits for the smallest stockpile.

**Fig 3.**
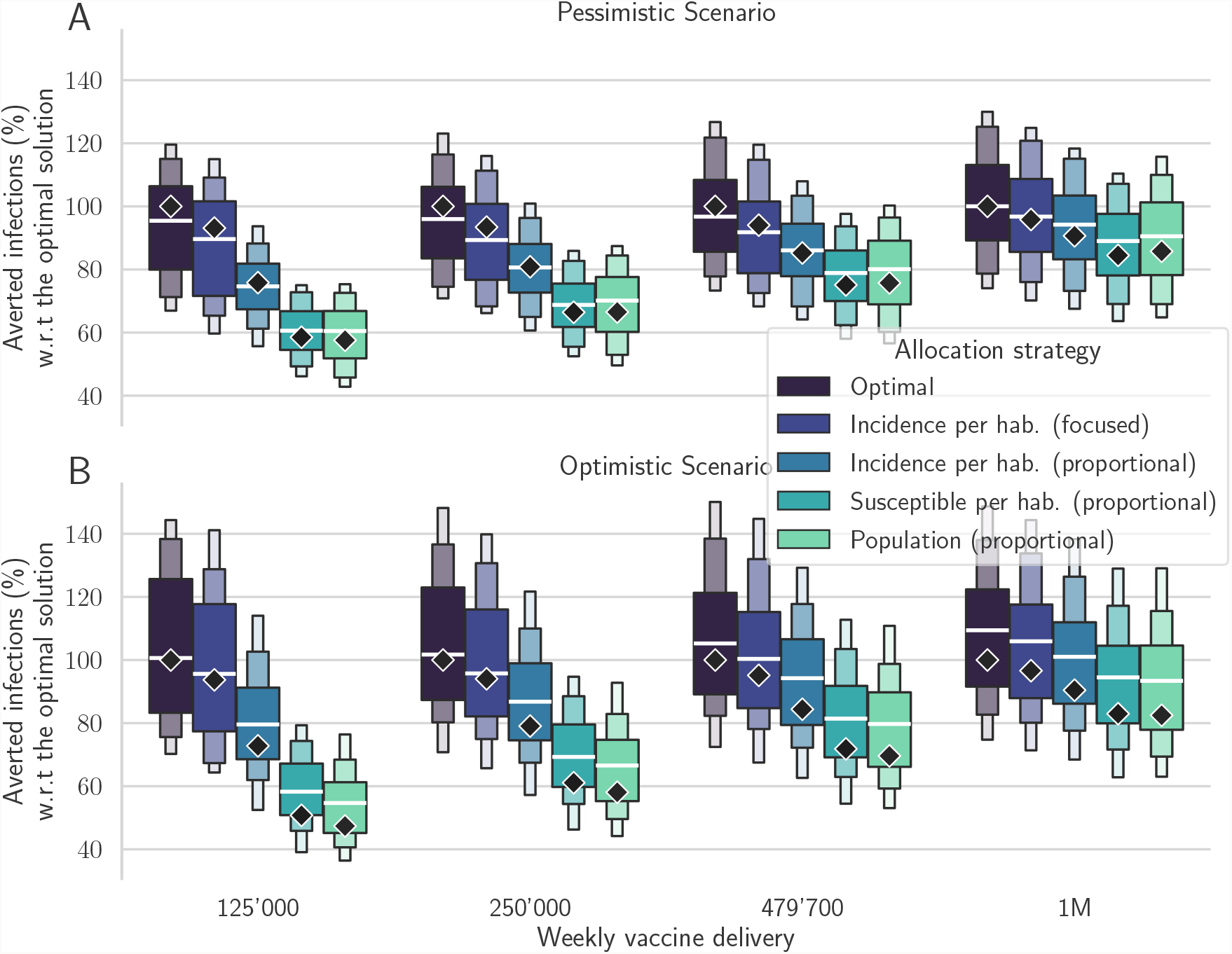
Comparison between different vaccine allocation strategies. Percentage of averted infections per vaccine doses from January 4, 2021 to April 4, 2021 resulting from province-scale vaccine allocation strategies for both the pessimistic (A) and the optimistic (B) scenarios based on the following vaccination strategies: the optimal solution, proportional to the province population, proportional to the susceptible individuals, proportional to the projected incidence, and focused on the provinces with the largest weekly incidence (see color codes in the legend). We optimize the vaccine allocation for the reference trajectory (the median trajectory in the model projections, indicated as diamonds in the figure), and assess the performance of the computed vaccination strategy over the whole posterior of trajectories (boxen plots). For each projection scenario, results are normalized by the number of averted infections in the reference solution (see Table 1 for the absolute values). Results for alternative scenarios and vaccination strategies are shown in SI, Figure S9.

In the pessimistic scenario (see Figure 3A), when 479’700 doses are available each week, the averted infections associated with the optimal strategy in the reference projection are 0.272 per dose: 24.6 % more compared to the strategies based on population or susceptible distributions (0.205 averted infections per dose), and more than 14 % higher compared to the strategy based on the projected incidence per inhabitant (0.232 averted infections per dose), while only 4% higher than the focused incidence strategy. These differences are smaller but still significant when increasing the weekly stockpile deliveries up to 1M doses; similar results are obtained also for the optimistic transmission scenario (Figure 3B).

We recall that the optimal control strategy considered here is computed for a reference model trajectory, which is the median of an ensemble of 100 realizations. To further investigate the effectiveness of the optimal solution, we apply it to a subset of trajectories randomly sampled from the ensemble. The box plots in Figure 3 display the main quantiles of the averted infections computed for the ensemble of trajectories. We observe that the optimal strategy still yields better results than the ensemble of projections related to the other strategies, thus suggesting that the computed solution is robust even under the presence of perturbations in the forecasts of the epidemic dynamics. More importantly, for each realization of the ensemble and for each projection scenario, the optimal strategy systematically averts more infections than any of the other control strategies.

Our results suggest that it is possible to considerably increase the impact of vaccination campaigns by optimizing the vaccine allocation in space and time. For this task, optimal control provides the best possible strategy and sets a benchmark for the theoretical potential of a vaccination campaign.

### Analysis of the optimal vaccine allocation

The optimal vaccine allocation obtained as the solution of the optimal control framework is complex to analyze, and we ought to do that by unraveling the mechanism behind its performance. The strategy must obey the imposed logistic and supply constraints: 1) The vaccine stockpile is replenished every Monday by a fixed amount of doses (e.g., 479’700 doses in the baseline scenario), and 2) the maximum possible distribution capacity per province is limited, proportionally to the node population, so that the number of doses distributed across the country can be of 0.5M per day at maximum (more details in SI Figure S2).

We display the optimal vaccination course in time for the pessimistic, 479’700 doses/week scenario in Figure 4. We observe that the optimal allocation respects the two constraints on distribution (Figure 4, A) and supply (Figure 4, B). We observe that no province is vaccinated at the maximum possible rate during the whole campaign, and that provinces display a variety of vaccination patterns. We also note that the vaccines received every Monday are always distributed during the following week, but that the rate of delivery on a national level increases with time (Figure 4, B). Surprisingly, the optimal solution favors precise targeting over speed of delivery, in order to allocate more doses to those provinces where the impact of vaccines on the whole system is projected to be higher. Hence, in order to control infections, precise targeting may trump delivery speed.

**Fig 4.**
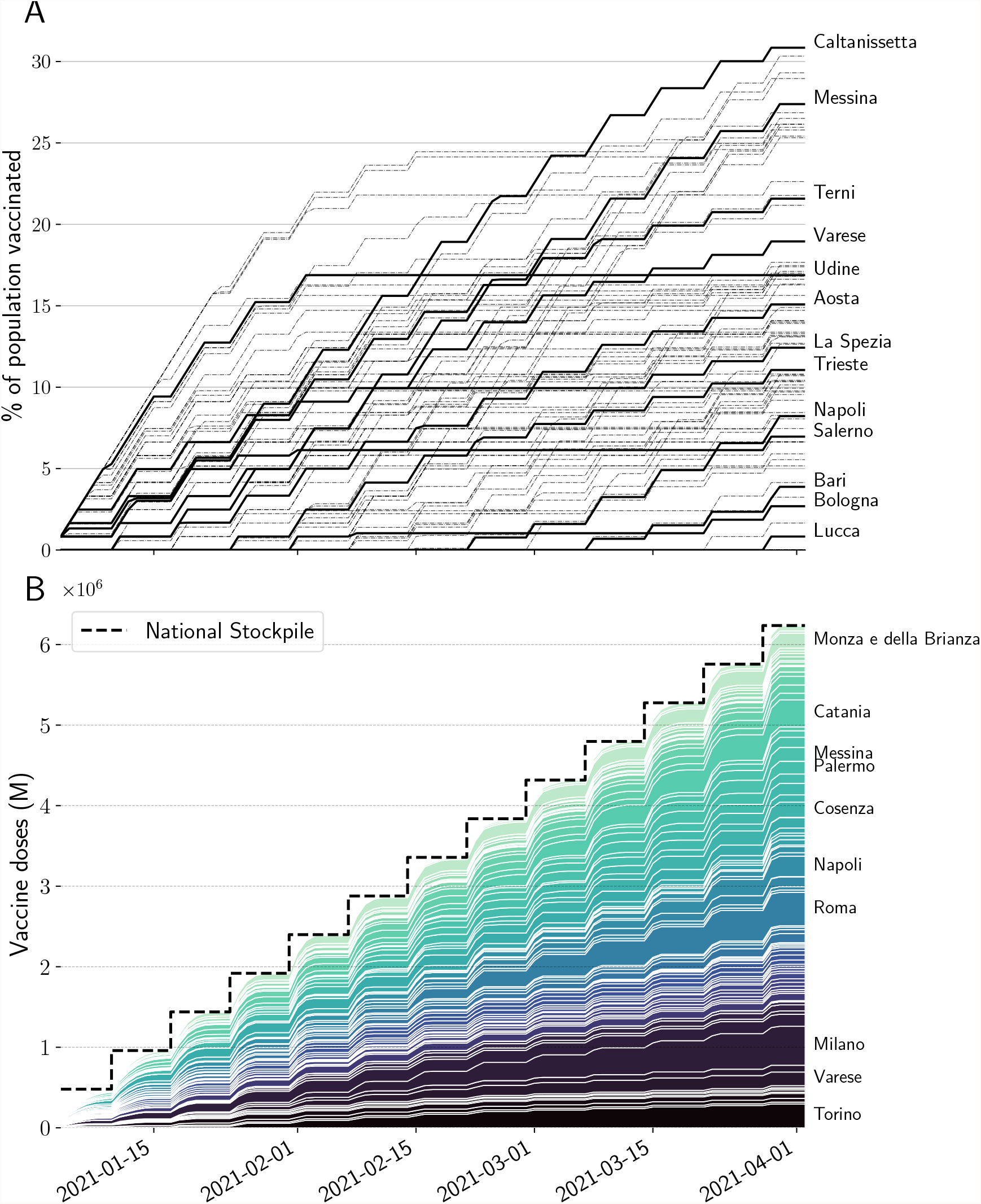
Optimal vaccine allocation for the baseline, pessimistic scenario. (A) Cumulative proportion of vaccine doses administered in the 107 provinces, (some of which are highlighted). The local distribution rate is limited by a rate that is proportional to population. This logistic constraint is visualized here as the maximum slope, equal for every province. (B) Stacked cumulative absolute number of vaccines in the 107 provinces of Italy. The national stockpile is shown in black, and is replenished every week (say on Mondays) with 479’700 doses. We display the name of the provinces with a final allocation of more than 150’000 doses.

Furthermore, we observe in the optimal solution that every time a province is vaccinated, the rate of vaccination is equal to the maximum rate allowed by the local logistic constraint, as it is the case for any focused alternative vaccination strategies. In Figure 5, one can already see by visual inspection that the optimal allocation distributes most of the available doses on a few provinces with high incidence. These provinces are neither the most connected nor the most populous in Italy. The optimal strategy makes then use of the information on the network connectivity to fine-tune the allocation, and deploys the vaccination on more provinces than the incidence-based strategy.

**Fig 5.**
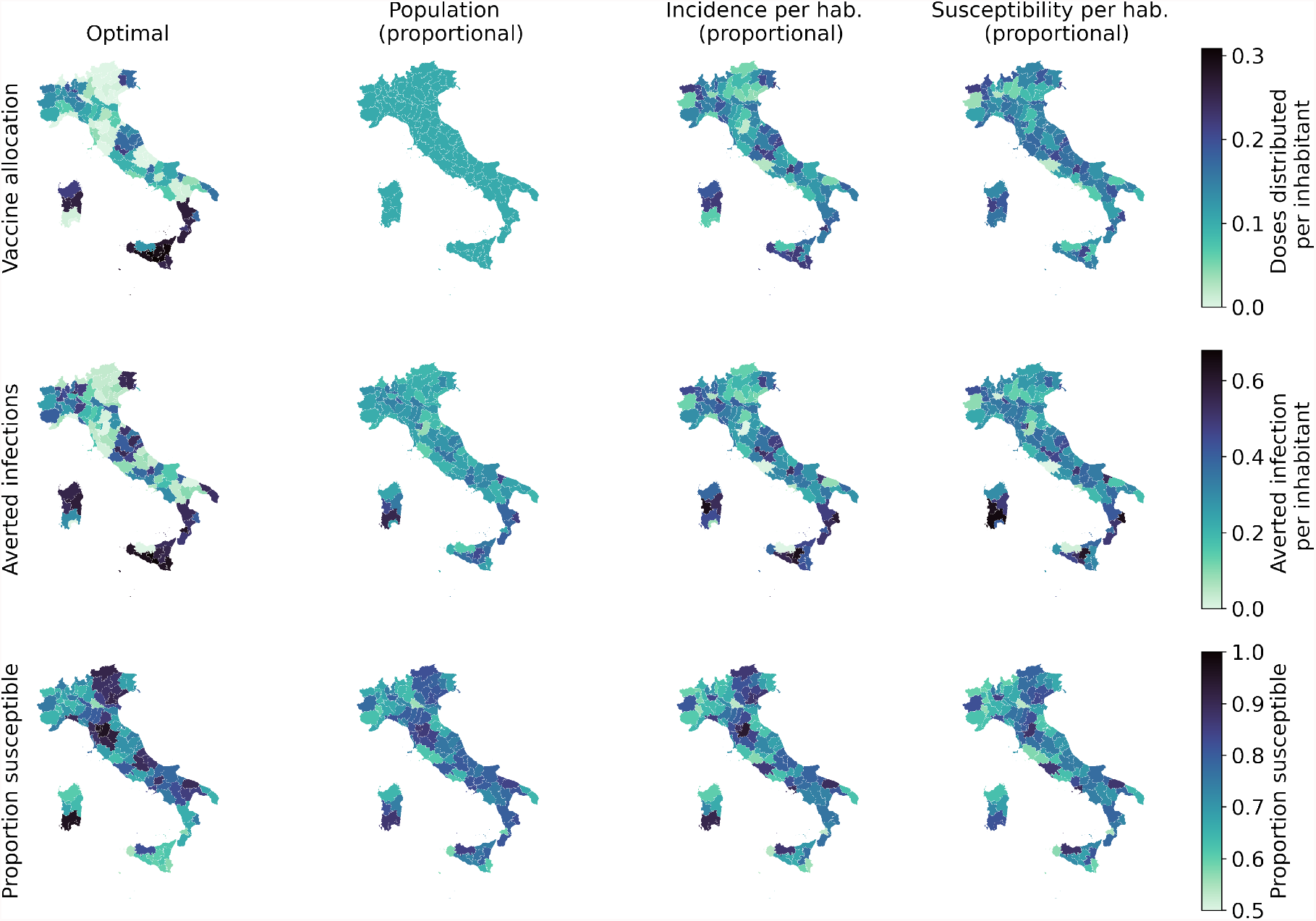
Spatial distribution patterns for the optimal allocation (left) and alternative strategies based on population, incidence and susceptibility (additional alternative strategies are presented in SI). We show, for each province and strategy, the proportion of vaccinated individuals (top), the number of averted infections per inhabitant with respect to a no vaccination baseline (middle), and the proportion of individuals who are still susceptible at the end of the control horizon (bottom).

To further investigate these patterns, in Figure 6A we display the number of administered doses vs the incidence projected without vaccines (the proxy variables leading to the second-best control performance), both normalized according to the resident population in each province. We observe an allocation pattern whereby provinces with a higher incidence receive more vaccines. However, the allocation is non-linear with respect to the projected incidence, suggesting that to better control the epidemic, the optimal allocation strategy takes into account other factors such as the importance of each province within the mobility network, as well as the proportion of susceptibles. When the weekly stockpile delivery is increased, as shown in Figure 6B, this pattern shifts to the right while remaining qualitatively consistent. Hence, the optimal allocation strategy is robust with respect to the overall vaccine availability constraint, and the same nodes are prioritized. We provide scatter plots with other covariates in SI (Figures S10–S11).

**Fig 6.**
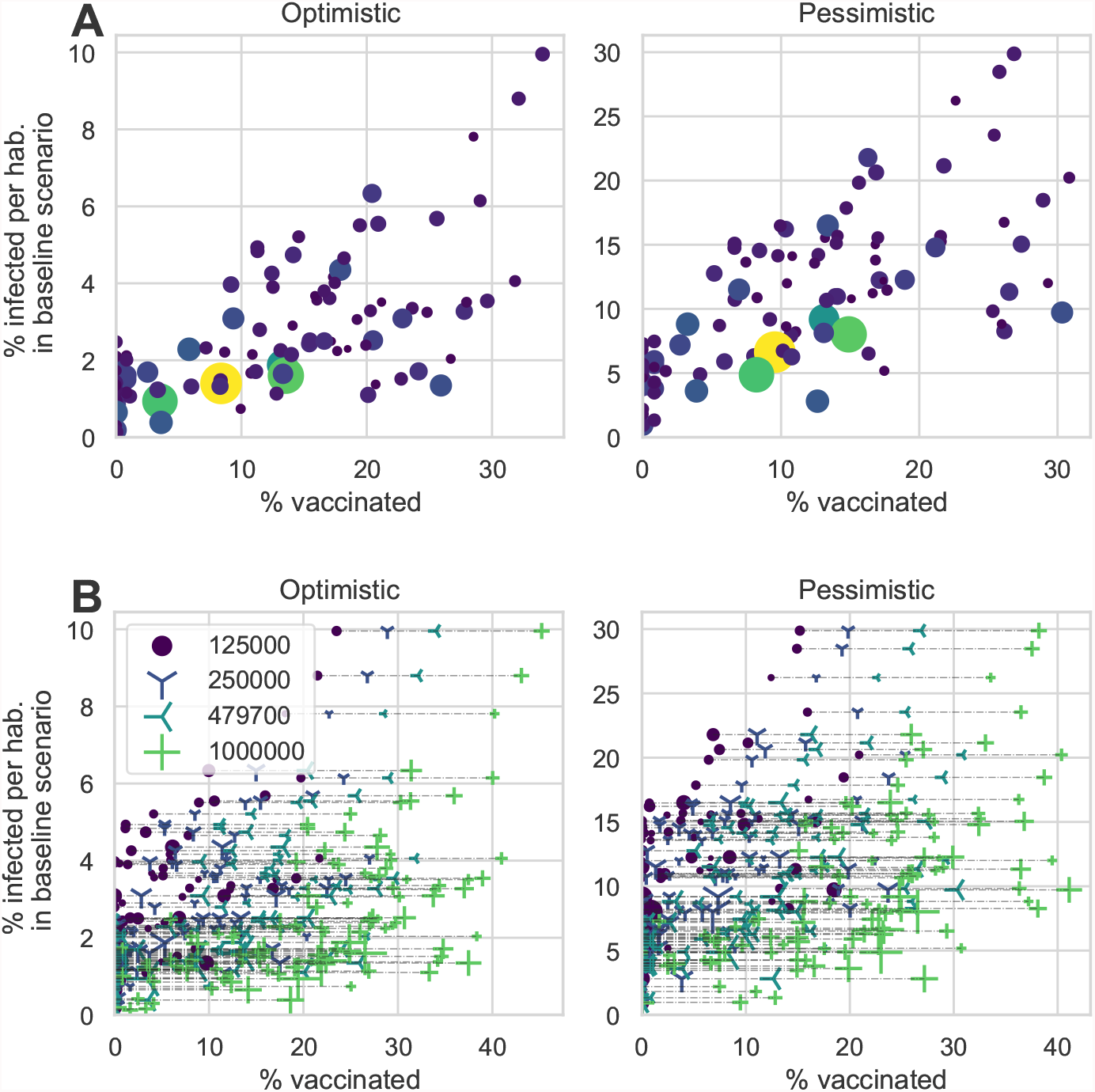
Analysis of the optimal solution. (A) Vaccinated population according to the optimal strategy against the projected incidence without vaccination, both normalized by provincial population size and considering the scenario with a weekly stockpile delivery of 479’700 doses. (B) Same as in A, but considering all four scenarios of weekly stockpile replenishment. Each dot represents a province, and the dot size is proportional to the population.

## Discussion and Conclusion

Without any constraint on supply, each country would vaccinate its population as fast as possible according to the available infrastructure. However limitations in vaccine supply and rate of delivery are a reality for every country, hence the available doses should be deployed in space and time following a fair and effective strategy.

In stockpile-limited settings, like most current vaccination campaigns worldwide, careful allocation may significantly increase the number of averted infections and deaths. The goal is to distribute the vaccines where they have the strongest beneficial impact on the dynamics of the epidemic. However, deriving an algorithm capable of computing spatially optimal allocation strategies in real, heterogeneous settings is far from trivial and our approach is, to the best of our knowledge, the first attempt in this direction.

We developed a novel optimal control framework that delivers the best vaccination strategy under constraints on supply and logistics. This allows us to compute the allocation strategy that maximizes the number of averted infections during a projection of the COVID-19 epidemic in Italy from January 4, 2021 to April 4, 2021. Our results show that the optimal strategy has a complex structure that mainly reflects the projected incidence of each province, but also takes into account the spatial connectivity provided by the mobility network and the landscape of acquired population immunity. Although the reason why this strategy is optimal is not immediately intuitive, our simulations clearly outline its better overall performances over other, more straightforward strategies. This comparison suggests that the simplicity underlying intuitive vaccination strategies may undermine their effectiveness, and calls for complementing these simple approaches with rigorous and objective mathematical tools, like optimal control, that allow a full account of the complexity of the problem.

With the present work, we show that it is possible to solve optimal control problems for spatially explicit dynamical models of infectious diseases at a national scale, thus overcoming the computational limitations that, up to now, precluded this kind of applications. The proposed framework can account for any compartmental epidemic model, with up to hundreds of connected spatial nodes. Supply and logistic constraints can be adapted to the actual landscape of decisions faced by the stakeholders, such as no/reduced vaccine delivery on weekends, or the need for fairness in vaccine distribution, e.g. by ensuring that each region receives at least a fixed fraction of the available vaccines. This is especially important as in our optimal allocation scenarios some provinces receive no vaccine at all. Moreover, the optimization can be carried for single-dose vaccines, as done here, or for two-dose vaccines, where one could potentially optimize the time between the first and second dose (and if a second dose should be administered at all), clearly also considering the intervals recommended by the health authorities.

Our method is obviously not devoid of limitations. The main one is that the optimal vaccination strategy strongly depends on the projection of the underlying epidemiological model. These projections are subject to several sources of uncertainty, especially for long term horizons, for example due to model design and calibration [47], the generation of baseline transmission scenarios, and unforeseen events that may change the course of the epidemic (such as the importation of cases, the emergence of new virus variants, changes in disease awareness or social distancing policies). The optimal vaccination strategy is thus reliable only if the projections given by the underlying model dynamics are sufficiently accurate. A successful approach developed by the automatic control community to tackle that issue, named Model Predictive Control [48], consists in compensating the performance losses expected over long horizons by constantly adapting the optimal strategy. In this context, Model Predictive Control might be implemented using the following steps: (a) at the beginning of each week, the state of the system is estimated by using newly acquired epidemiological data; (b) the optimization problem is solved over a fixed prediction horizon using the estimated state as initial condition; (c) the optimal strategy for the first week is applied and, as soon as the next week starts, these steps are repeated starting from (a). This method corrects the model inaccuracies by constantly resetting the initial state to the estimated one. Additionally, constraints may be updated to account for unexpected deliveries or new orders. Future work will aim at further evaluating the benefits of implementing this scheme for the design of optimal vaccination strategies. Further refinement could be obtained accounting for the whole uncertainty range while optimizing instead of only median evolution of the epidemic using methods from robust or stochastic model predictive control. This would improved the proposed framework to provide reasonable performance in every case. At the moment, this is not computationally tractable. The sensitivity analysis provided in SI at least shows that the optimal allocation strategy does not perform comparatively poorly if the model projections are inaccurate.

The epidemiological model underlying our control optimization has known validity and limitations [8, 9]. An additional limitation of the model for the specific scopes of this work is that it does not account explicitly for risk-based classes, and thus does not account for the heterogeneities that may result from the demography of the population, as well as from the age-related transmission and clinical characteristics associated with COVID-19. While surely limiting for operational use of the tools, we note that the scope of this paper is to provide a proof-of-concept of the relevance of spatial effects, which have not been addressed so far in the literature. To that end, we are confident that our results support the relevance of the research question posed. Our framework can anyway be extended to optimize across both spatial and risk heterogeneities, provided the accessibility to the computational and storage capacities that the solution of such optimal problem requires.

A counter-factual assumption in this work is that we consider a one-dose vaccine with full and instantaneous efficacy against transmission. At the time of development, the details about COVID-19 vaccines were not released, and this hypothesis allowed us to demonstrate our framework in a simple setting. Our framework can be further extended to account also for the simultaneous deployment of different vaccine types, some of which may require the administration of two doses. This extension too is subject of ongoing research, in particular to extend the modeling tools described here to accommodate the peculiarities of each authorized vaccine candidate while designing effective spatiotemporal deployment strategies.

In conclusion, in this work we have optimized vaccine allocation at country scale on different scenarios of epidemic transmission and vaccine availability. Using a data assimilation scheme, we updated a spatially explicit compartmental model that had already been successfully used to describe the COVID-19 pandemic in Italy. To this aim, we have discretized, transformed and simplified the model and constructed a pipeline to perform large-scale nonlinear optimization on vaccine allocation, subject to stockpile and logistic constraints. Solving this problem yielded a complex solution that outperforms other strategies by a significant margin and proves robust across posterior realizations of the underlying model. As such, beside inherent limitations, it provides a benchmark against which other, possibly simpler vaccine rollout strategies can be usefully compared.

## Supporting information

Supplemental info: additional methods and results

## Data Availability

The full framework and analysis code to generate the figures is available here: https://github.com/jcblemai/COVID-19_italy-vaccination-oc

## Acknowledgments

JCL and AR acknowledge funding from the Swiss National Science Foundation via the project “Optimal control of intervention strategies for waterborne disease epidemics” (200021–172578). AR, EB and DP acknowledge funding from Fondazione Cassa di Risparmio di Padova e Rovigo (IT) through its grant 55722. DP, EB, LM, RC, MZ and JCL thanks the FISR founding for the project EPIDOC (Epidemiological Data assimilation and Optimal Control for short-term forecasting and emergency management of COVID-19 in Italy), FISR-2020IP-04249. JCL acknowledge Nicolas Richart for help on scaling up computations on the EPFL high-performance computing clusters.

## Supporting Informations

Supporting information file si.pdf presents detailed methods and further results analysis.

