## Supplemental info: additional methods and results for "Optimal control of the spatial allocation of COVID-19 vaccines: Italy as a case study"

### Supporting information for Optimal control of the spatial allocation of COVID-19 vaccines: Italy as a case study

#### Optimal control problem

The proposed framework is based on two disease transmission models, a full model and a simplified one used for control:

- The full model is the COVID-19 model designed in [1, 2]. This model is based on ODEs, it includes full connectivity fluxes among nodes which are estimated from mobility data, and it is implemented in MATLAB using the explicit Runge-Kutta (4,5) integration scheme with adaptive temporal step (ode45). Using data assimilation, we obtain the temporal variations of the regional transmission parameters of this model.
- The simplified model used for optimal control is an approximation of the above model, integrated using an explicit Runge-Kutta 4 method with fixed stepsize. We simplified the problem by limiting the connectivity to the largest mobility fluxes (see Figure 1 B) and optimizing only one realization of the posterior. This model is implemented in Python with the CasADi library.

The simplifications introduced in the second model are necessary in order to solve the optimal control problem (OCP) in a reasonable time. To adapt our framework to another model/country, one would need to update the "true" model to a suitable candidate (which could be a stochastic model, a Hidden Markov model, or any other kind) and design a tractable approximation of this new model to be solved by optimal control.

In order to evaluate the effectiveness of our approach, we first compute the optimal vaccination course that minimizes the objective based on the simplified model. Then, we assess this strategy and the alternative ones on the full model, for different posterior realisations. If the simplified model is sufficiently accurate, the performance loss is small and the proposed strategy outperforms simpler strategies, as shown in our simulation results.

It the subsections below, we first detail the full COVID-19 model, then we describe the optimal control framework and the simplifications we introduced to bring the problem to a tractable form.

##### COVID-19 model

The optimal control framework may be used with any compartmental SARS-CoV-2 transmission model that can be approximated by ordinary differential equations. To demonstrate its usefulness, we consider a complex epidemiological model developed in previous work to describe the first wave of COVID-19 infections in Italy [1, 2]. The model subdivides the Italian population into the 107 Italian provinces, and connects them on the bases of human mobility fluxes. In each province, the human population is further subdivided according to its infection status into the epidemiological compartments of susceptible $S$, exposed $E$, pre-symptomatic $P$ (incubating infectious), symptomatic infectious $I$, asymptomatic infectious $A$, quarantined $Q$, hospitalized $H$, recovered $R$, dead $D$, and vaccinated $V$. The possible transitions between these compartments are shown in Figure 1A. Individuals in compartments $P$, $A$ and $I$ are infectious and contribute differently to the force of infection, driving susceptible $S$ into exposed individuals $E$.

The COVID-19 transmission dynamics are described by the following set of ordinary differential equations in each node $i$:

$$
\begin{aligned}
\dot{S}_i &= -\lambda_i(t)S_i - r_i^v(t)S_i \\
\dot{E}_i &= \lambda_i(t)S_i - (\delta^E + r_i^v(t))E_i \\
\dot{P}_i &= \delta^E E_i - (\delta^P + r_i^v(t))P_i \\
\dot{I}_i &= \sigma\delta^P P_i - (\gamma^I + \eta)I_i \\
\dot{A}_i &= (1-\sigma)\delta^P P_i - (\gamma^A + r_i^v(t))A_i \\
\dot{Q}_i &= \zeta\eta I_i - \gamma^Q Q \\
\dot{H}_i &= (1-\zeta)\eta I_i - (\gamma^H + \alpha^H)H \\
\dot{R}_i &= \gamma^I I_i + \gamma^A A_i + \gamma^H H_i + \gamma^Q Q_i - r_i^v(t)R_i \\
\dot{V}_i &= r_i^v(t) \cdot (S_i + E_i + P_i + A_i + R_i).
\end{aligned}
\tag{S1}
$$

Let $N_i$ be the population of province $i$. Susceptible individuals get exposed to the pathogen at rate $\lambda_i(t)$, corresponding to the force of infection for community $i$, thus becoming latently infected (but not infectious yet). Exposed individuals transition to the post-latent, infectious stage at rate $\delta^E$. Post-latent individuals progress to the next infectious classes at rate $\delta^P$, developing an infection that can be either symptomatic—with probability $\sigma$—or asymptomatic—with probability $1-\sigma$. Symptomatic infectious individuals recover from infection at rate $\gamma^I$ and may seek treatment at rate $\eta$. Asymptomatic individuals recover at rate $\gamma^A$. Infected individuals who sought treatment are either hospitalized (rate $1-\zeta$) or quarantined (rate $\zeta$) at home and are considered to be effectively removed from the community, thus not contributing to disease transmission. Individuals who recover from the infection are assumed to have long-lasting immunity to reinfection at the timescale studied, but possible loss of immunity can be easily included in the model. Hospitalized individuals die at rate $\alpha_H$ and recover at rate $\gamma^H$.

Individuals in compartments $S, E, P, A, R$ might receive vaccine doses. If the chosen strategy allocates $v_i(t)$ doses in node $i$ at time t, the vaccination rate is

$$
r_i^v(t) = \frac{v_i(t)}{S_i(t) + E_i(t) + P_i(t) + A_i(t) + R_i(t)} \tag{S2}
$$

Vaccinated individuals are moved at rate $r_i^v(t)$ from their original compartments to compartment $V$, where they do not contribute to the infection anymore.

The force of infection of the full model is specified as in Gatto et *al.* [1], Bertuzzo et *al.* [2]. In addition to province's local dynamics, it also considers that local susceptibles may enter in contact with infected individuals that are traveling, and oppositely, susceptible commuters may become infected through contact with local infected. The force of infection of the OCP model is slightly simplified, and detailed thereafter.

We split the force of infection $\lambda_i(t)$ as the sum of the local force of infection $\lambda_i^L(t)$, from infected in node $i$ and a mobility-driven force of infection from the network $\lambda_i^N(t)$, hence $\lambda_i(t) = \lambda_i^L(t) + \lambda_i^M(t)$. We observe while running our model that $\lambda_i^M(t) \ll \lambda_i^L(t)$. Hence this artificial separation will be exploited when simplifying the model for the OCP. As described below, we update $\lambda_i^M(t)$ every day whereas $\lambda_i^L(t)$ is updated at each integration step.

As for the formulations of the force of infection, we recall here the formulas designed by [1, 2]. The local force of infection reads:

$$\lambda_i^L(t) = C_{i,i}(t)\beta_0\beta_i(t) \cdot \frac{C_{i,i}(t)(P_i + \epsilon_A A_i) + \epsilon_I I_i}{C_{i,i} \cdot (S_i + E_i + P_i + R_i + A_i + V_i) + I_i}, \tag{S3}$$

and the influence of other provinces on province $i$ is written as:

$$\lambda_i^M(t) = \sum_{m, m\neq i} \left( C_{i,m}(t) \cdot \frac{\sum_{n, n\neq m} [C_{n,m}(t) \cdot \beta_0\beta_n(t)(P_n + \epsilon_A A_n)] + \epsilon_I \beta_0 \beta_m(t) I_m}{\sum_{l, l\neq m} [C_{l,m}(t) \cdot (S_l + E_l + P_l + R_l + A_l + V_l)] + I_m} \right), \tag{S4}$$

where $\beta_0$ is the baseline transmission rate, and the parameters $\epsilon_A$ and $\epsilon_I$ represent the reduction of transmission respectively for asymptomatic and symptomatic individuals with respect to pre-symptomatic individual transmissions. Matrix $C(t)$ accounts for mobility: each element $C_{i,j}(t)$ of the matrix ($i \neq j$) represents the proportion of contacts among individuals moving from $i$ to $j$, while the diagonal elements $C_{i,i}(t)$ are the proportions of contact for individuals in node $i$ that do not move. With more details (see [1]):

$$C_{i,j} = \begin{cases} (1-p_i) + (1-r)p_i + rp_iq_{ij} & \text{if } i = j \\ rp_iq_{ij} & \text{otherwise} \end{cases} \tag{S5}$$

where $p_i$ is the fraction of mobile people commuting from node $i$ to the other nodes, $q_{ij}$ is the fraction of mobile people between nodes $i$ and $j$, and $r$ is an additional parameter describing the fraction of time spent traveling (here set to 0.5 days). Fractions $p_i$ and $q_{ij}$ were estimated in [1]) from the commuting mobility assessment of Italy performed in 2011 by the Italian National Institute of Statistics (ISTAT, data available at `https://www.istat.it/it/archivio/139381`).

Here we assume that the initial local fraction of mobile individuals $p_i$, in the following indicated with $p_{i0}$, changes during the simulation due to the mobility restrictions imposed by the national and regional governments during the epidemic. We adopted the mobility trends for places of work estimated in the Google COVID-19 Community Mobility Reports [3] as a proxy for the reduction in mobile individuals in a province $i$ and day $t$,

$$p_i(t) = p_{i,0}(1 + g_i(t)/100)$$

where $g_i(t)$ is the percentage of change in mobility in province $i$ and day $t$ with respect to the mobility at the beginning of February 2020, as reported by the Google data (data shown in Figure S7).

The model further exploits the Google estimates in mobility reduction as a proxy of changes in individual awareness and social distancing. This is represented in parameter $\beta_i(t)$, a spatially distributed and time-varying parameter describing site- and time-specific variations in transmission due to non-pharmaceutical interventions or other exogenous factors like variants. At a given day $t$, we pose

$$\beta_i(t) = (1 + g_i(t)/100)\phi_i(t) \tag{S6}$$

where parameter $\phi_i(t)$ changes at the regional level and is calibrated in the data assimilation procedure in order to fit the hospitalization data (see section Data assimilation and model parameters, Figure S7)

The objective for our model is to minimize the total incidence of infections, i.e., $\int_{t_i}^{t_f} \sum_i \lambda_i(t) S_i$. Note that for the present model, this is equivalent to optimizing the total deaths or hospital admissions, as without risk-classes the sizes of these two compartments are proportional to each other.

#### Age structure

For the calibration and assimilation procedure only, a post-processing algorithm subdivides the modelled number of daily new exposed individuals, $E_i(t)$, into five age classes: $a_1 = 0-19$, $a_2 = 20-39$,$a_3 = 30-59$,$a_4 = 60-79$;$a_5 = 80+$.

Let $S_i^{(j)}(t)$ and $E_i^{(j)}(t)$ be the number of susceptible and new exposed individuals in node $i$ ($i = 1, \ldots, n$) and age class $j$ ($j = 1, \ldots, 5$), at time $t$, with the property that $\sum_j S_i^{(j)}(t) = S_i(t)$ and $\sum_j S_i^{(j)}(t) = E_i(t)$.

The new exposed individuals per age class are a fraction $p_i^{(j)}(t)$ of the total new exposed:

$$E_i^{(j)}(t) = p_i^{(j)}(t)E_i(t) \tag{S7}$$

where $\sum_j p_i^{(j)}(t) = 1$. This fraction takes into account the possible different exposure, susceptibility, and number of contacts that characterizes the age class, indicated with $c_i^{(j)}$, and also the number of susceptible individuals still present in the node for that age class, $S_i^{(j)}(t)$:

$$p_i^{(j)}(t) = c_i^{(j)} \frac{S_i^{(j)}(t)}{S_i(t)} \tag{S8}$$

We estimate the coefficients $c_i^{(j)}$ on the basis of epidemiological data. Before the beginning of the vaccination campaign (December 2020, see [4]) the total reported cases of COVID-19 in Italy where subdivided as in the following: $p^{(1)} = 12.13\%$, $p^{(2)} = 24.23\%$, $p^{(2)} = 33.92\%$, $p^{(2)} = 19.58\%$, $p^{(2)} = 10.14\%$.

Assuming that the depletion of susceptibles and reporting issues have a negligible impact on these cumulative statistics, we compute the coefficients $c_i^{(j)}$ as follows:

$$c_i^{(j)} = p^{(j)} \frac{N_i}{N_i^{(j)}}$$

where $N_i^{(j)}$ is the population in node $i$ and age class $j$ (data from [5] ).

The post-processing algorithm is initialized with $S_i^{(j)}(0) = N_i^{(j)}$. In a day $t$, assuming to know $S_i^{(j)}(t)$, the algorithm follows these steps:

- eq. (S8) evaluates the probabilities $p_i^{(j)}(t)$, which are normalized if needed;
- the new exposed $E_i^{(j)}(t)$ are subdivided per age class using (S7);
- the number susceptibles are updated by removing the new exposed and the vaccinated susceptibles in that node and age class:

$$S_i^{(j)}(t+1) = S_i^{(j)}(t) - E_i^{(j)}(t) - V_i^{(j)}(t)$$

- The number of deaths $D_i^{(j)}(t+\tau)$ per age class are computed from the exposures through an age-class specific case fatality rate $\mu^j$ (equal per each node):

$$D_i^{(j)}(t+\tau) = \mu^j E_i^{(j)}(t+\tau)$$

  where $\tau = 14$ days is the average time between exposure and death (see [6]), and $\mu^1 = 0.01\%$, $\mu^2 = 0.04\%$ $\mu^3 = 0.43\%$, $\mu^4 = 6.06\%$, $\mu^5 = 20.83\%$ (data from [4] )

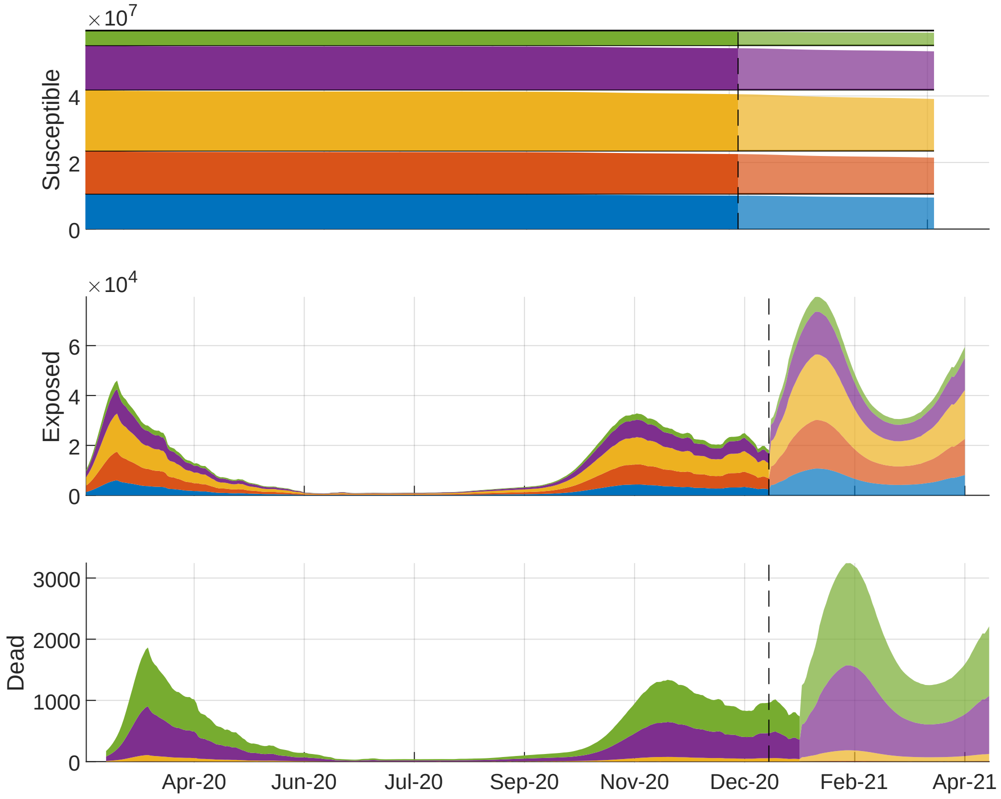


**Fig S1. Age-stratified outputs.** Results of the post-processing algorithm for the computation of susceptible (panel A), incidence (panel B) and deaths (panel C) among the five considered age classes. The algorithm provides results at the province level, which are here aggregated at the national level (see SI, section on age structure.)

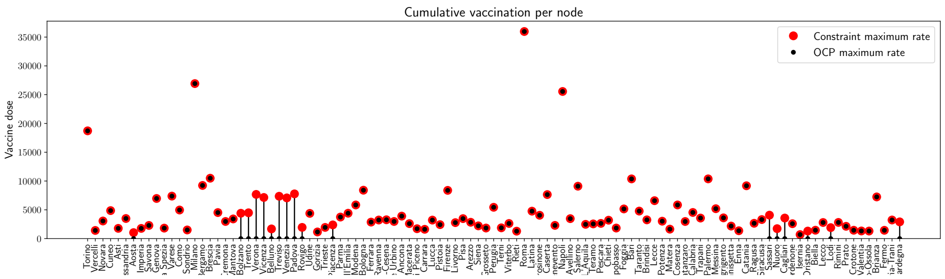


**Fig S2. Local maximum vaccination rate $v_i^{\max}$ for each province.** This logistic constraint bounds the maximum number of vaccines to 0.5M of doses per day, with a local rate that is proportional to the node population. Here we show the maximum vaccination rate for each province (the constraint the solution has to comply with), in red, and the maximum rate prescribed by the optimal solution while simulating the pessimistic scenario with a stockpile delivery of 479’700 doses, in black. The optimal solution uses the maximal capacity of the logistic network, while respecting the constraint defined.

#### Optimal control

We lump the epidemiological compartments of each node $i$ in variable $x_i(t) = (S_i(t), E_i(t), P_i(t), I_i(t), A_i(t), Q_i(t), H_i(t), R_i(t), V_i(t))$ and we define $v_i(t)$ as our control variable, representing the number of vaccines administered in node $i$ at time $t$. We express the dynamics of the epidemiological model (Equation (S1)) as an ordinary differential equation in each province $i$:

$$\dot{x}_i(t) = F_i(x_i(t), v_i(t), m_i(t), t), \tag{S9}$$

where $m_i(t)$ carries the contribution of other provinces to the force of infection of node $i$. For simplicity, we drop the time dependence in the equations below, and we define the state and control variables for the full system as

$$x = (x_1, \dots, x_n), \qquad v = (v_1, \dots, v_n),$$

where $n$ is the number of spatial node considered ($n = 107$). The global dynamics for all provinces are denoted:

$$F(x, v) = (F_1(x_1, v_1, m_1), \dots, F_n(x_n, v_n, m_n)).$$

The coupled force of infection in node $i$ is denoted $\lambda_i$. We define the cost function as the sum of total incidence of infections (transitions $S_i \longrightarrow E_i$) for every node $i$, i.e.,

$$L(x, v) = \sum_{i=1}^{n} \lambda_i S_i.$$

For the sake of generality, we introduce the terminal cost $M$, which can be used to ensure that we leave the system in a proper state instead of optimizing for short-term gain. Since properly designing the terminal cost could require a long analysis, for simplicity we do not use it in this work, hence $M(\cdot) = 0$.

Given our dynamical system with states $x$, controls $v$, and dynamics $F$, the OCP is:

$$\min_{v(\cdot)} \quad \int_0^T L(x(t), v(t)) \, \mathrm{d}t + M(x(T)) \tag{S10a}$$

$$\text{s.t.} \quad x(0) = \hat{x}_0, \tag{S10b}$$

$$\dot{x}(t) = F(x(t), v(t)), \qquad \forall\, t \in [0, T], \tag{S10c}$$

$$H(x(t), v(t)) \leq 0, \qquad \forall\, t \in [0, T], \tag{S10d}$$

where we aim at minimizing the cost function over the prediction horizon $T$, while enforcing the modeled SARS-CoV-2 transmission dynamics (Equations (S10b) and (S10c)). Moreover, constraints on vaccine availability and maximum vaccination rate are lumped in function $H$, which reads:

$$v_i(t) \geq 0, \qquad i \in \mathbb{I}_1^n, \tag{S11a}$$

$$\int_{t_\mathrm{d}}^{t_\mathrm{d}+1} v_i(t) \, \mathrm{d}t \leq v_i^{\max} \propto N_i, \qquad i \in \mathbb{I}_1^n,\ t_\mathrm{d} \in \mathbb{I}_0^T, \tag{S11b}$$

$$\int_0^t \sum_{i=1}^{n} v_i(t) \, \mathrm{d}t \leq D(t), \qquad \forall\, t \in [0, T], \tag{S11c}$$

where time is measured in days, and $\mathbb{I}_a^b$ is the set of all integers $a \leq k \leq b$. Equation (S11a) enforces that one can only distribute a non-negative amount of vaccine doses. Equation (S11b) states the logistic constraints, which limit the amount of individuals that can be vaccinated each day in each node to $v_i^{\max}$; here $t_\mathrm{d}$ is the time at which each day starts. We assume that the daily capacity of each province is proportional to the population size of each node $N_i$, because we assume a fair distribution of the sanitary infrastructure among provinces with regard to population, as shown in SI Figure S2. The stockpile is materialized by Equation (S11c), which ensures that the total vaccine allocation across every node does not exceed the total availability $D(t)$. The stockpile is replenished every Monday by the delivery of new vaccines, hence $D(t)$ is a staircase function.

We convert our problem formulation (S10) to a nonlinear programming problem using direct multiple shooting. Standard multiple shooting splits the time horizon $[0, T]$ using a time grid $t_0, \dots, t_N$, with $N + 1$ points and $t_0 = 0$, $t_N = T$. The control function is parameterized using basis functions with local support. Common choices are a uniform time grid, i.e., $t_{k+1} = t_k + \delta_t$ and a piecewise constant control function, i.e., $v(t) = v_k$, $t \in [t_k, t_{k+1}]$. The system dynamics are then discretized to obtain a discrete-time system

$$x_{k+1} = f(x_k, v_k),$$

satisfying $x_k = x(t_k)$ for all $k = 0, \ldots, N$. Moreover, the cost function is also discretized, to obtain

$$l(x_k, v_k) = \int_{t_k}^{t_k+1} L(x(t), v(t)).$$

We perform the discretization using numerical integration techniques (such as a fourth-order Runge-Kutta scheme, with 50 steps per days) to obtain a good approximation of the true trajectory and cost. Finally, the path constraints $H$ are relaxed and imposed at a finite amount of time instants, here coinciding with the time grid $t_0, \ldots, t_N$. We ought to observe that, since in our case the constraints only involve the controls, we are not introducing any approximation by enforcing these constraints only on this uniform grid. The OCP (S10) is then approximated by the nonlinear programming problem

$$\min_{x,v} \quad M(x_N) + \sum_{k=0}^{N-1} l(x_k, v_k) \tag{S12a}$$

$$\text{s.t.} \quad x_0 = \hat{x}_0 \tag{S12b}$$

$$x_{k+1} = f(x_k, v_k), \qquad k \in \mathbb{I}_0^{N-1}, \tag{S12c}$$

$$H(x_k, v_k), \qquad k \in \mathbb{I}_0^{N-1}. \tag{S12d}$$

In (S12), both the states $x = (x_0, \ldots, x_N)$ and the controls $v = (v_0, \ldots, v_{N-1})$ are defined as optimization variables, which is a distinguishing trait of multiple shooting as opposed to single shooting.

The main difficulty in solving (S12) in the context of this paper is the large dimension of the system and the nonlinearity of the model, which can pose severe issues to the numerical solvers. In the following, we will thus introduce a few simplifications, and we will verify through numerical simulations that these simplifications do not imply large errors in the solution of the OCP (see Figure S3).

We discretize the OCP using a uniform grid with sampling time $\delta t = 1$ day. We assume that (a) vaccinations are administered instantaneously at the beginning of each day, rather than with a constant rate over the whole day; (b) the force of infection associated with mobility is constant over each day; and (c) the weakest mobility links can be pruned. Thus, each node dynamics can be made independent of the other nodes dynamics by introducing an auxiliary control variable $z$ that is constrained to match the force of infection due to the other nodes at the beginning of each time interval. Then, the dynamics of the decoupled system in each node can be written as:

$$\begin{aligned} \dot{x}_i(t) &= F_i(x_i(t), z_{i,k}), && t \in [t_k, t_{k+1}] \\ x_i(t_k) &= x_{i,k} + g_i(v_{i,k}), && z_{i,k} = e_i(x). \end{aligned}$$

**Discussion on Simplification (a).** We ought to remark that, realistically, vaccinations will occur at least eight hours per day. Our assumption, while justified as a computationally convenient approximation of reality, is not a priori worse than assuming that vaccine administration takes place over the whole day. More refined approximations, while in principle possible, pose severe issues because of the nature of the system dynamics. While for most initial values the system dynamics can be easily simulated with time-continuous vaccinations, the system becomes stiff by construction once almost the entire population has been vaccinated. In this case, numerical integration errors can drive the size of some compartments to be negative, which violates the model assumptions and makes the result of the numerical integration meaningless. The main issue in this case is that the optimizer will exploit these inaccuracies in order to reduce the cost. Therefore, this issue is much more evident when solving optimal control problems than when simply simulating the system dynamics. We have investigated some simple approaches to tackle this issue, but no technique yielded satisfactory performances. It is our impression that ad-hoc integration strategies will be required in order to reliably simulate and optimize dynamics with continuous vaccination rates. While this will be the subject of future research, the results obtained with the current approximation have yielded sufficient accuracy.

**Discussion on Simplification (b).** This simplification has been proposed in [7] as an approach to solve distributed optimal control problems by means of multiple shooting. In the original version, the coupling variable $z$ is not necessarily piecewise constant, but rather piecewise polynomial. We have observed in simulations that, for this problem, the piecewise constant parametrization yielded sufficient accuracy.

We discretize the dynamics of each node using an explicit Runge-Kutta integrator of order four, with 50 integration steps per day. Alternative integrators such as explicit Euler, or implicit Runge-Kutta integrators, yielded similar results. Furthermore, in order to verify the accuracy of the integrator and the impact of the introduced simplifications on the solution accuracy, we simulated the system in open-loop, i.e. we applied the optimal control trajectory to the full model starting from the initial condition provided by the data assimilation scheme.

**Discussion on Simplification (c).** We sparsify the mobility matrix by pruning element below a threshold (see Figure S4). This operation reduces the number of connection between nodes. Also in this case, we verified through numerical simulations that the introduced simplification had a small impact on the prediction and control accuracy.

**Possible further improvements** Applying optimal control in open loop, i.e., solving the optimization problem once and applying the control input over the whole time interval, may lead to poor performance due to model inaccuracy and external perturbations. A common remedy consists in closing the loop by repeatedly solving the OCP by using the most updated information on the initial states. This is the principle behind Model Predictive Control (MPC) [8]. In this context, the state would be estimated on a daily, weekly, or monthly basis so as to solve again the OCP and correct the optimal strategy.

#### Implementation of the OCP

We implement the optimal control framework using the automatic differentiation framework CasADi [9], the interior-point solver ipopt [10], and the HSL ma86 large sparse symmetric indefinite solver [11]. The full framework and analysis code is available here:
`https://github.com/jcblemai/COVID-19_italy-vaccination-oc` (a zenodo DOI will be added after reviews).

Solving the OCP is both CPU and RAM intensive. For numerical computations, we used the Helvetios cluster the EPFL HPC facility (one problem per computing node, each equipped with 36 2.3 GHz cores and 192 GB RAM). On this cluster, it takes approximately four days to solve the large-scale OCP just presented. It should be possible to solve even larger problems with more RAM available.

### Data assimilation and model parameters

The local transmission rates computed as $\beta_0\beta_i(t) = \beta_0(1 + g_i(t)/100)\phi_i(t)$ (S6) are the main parameters governing the force of infection of the model and, thus, the daily exposed individuals. To

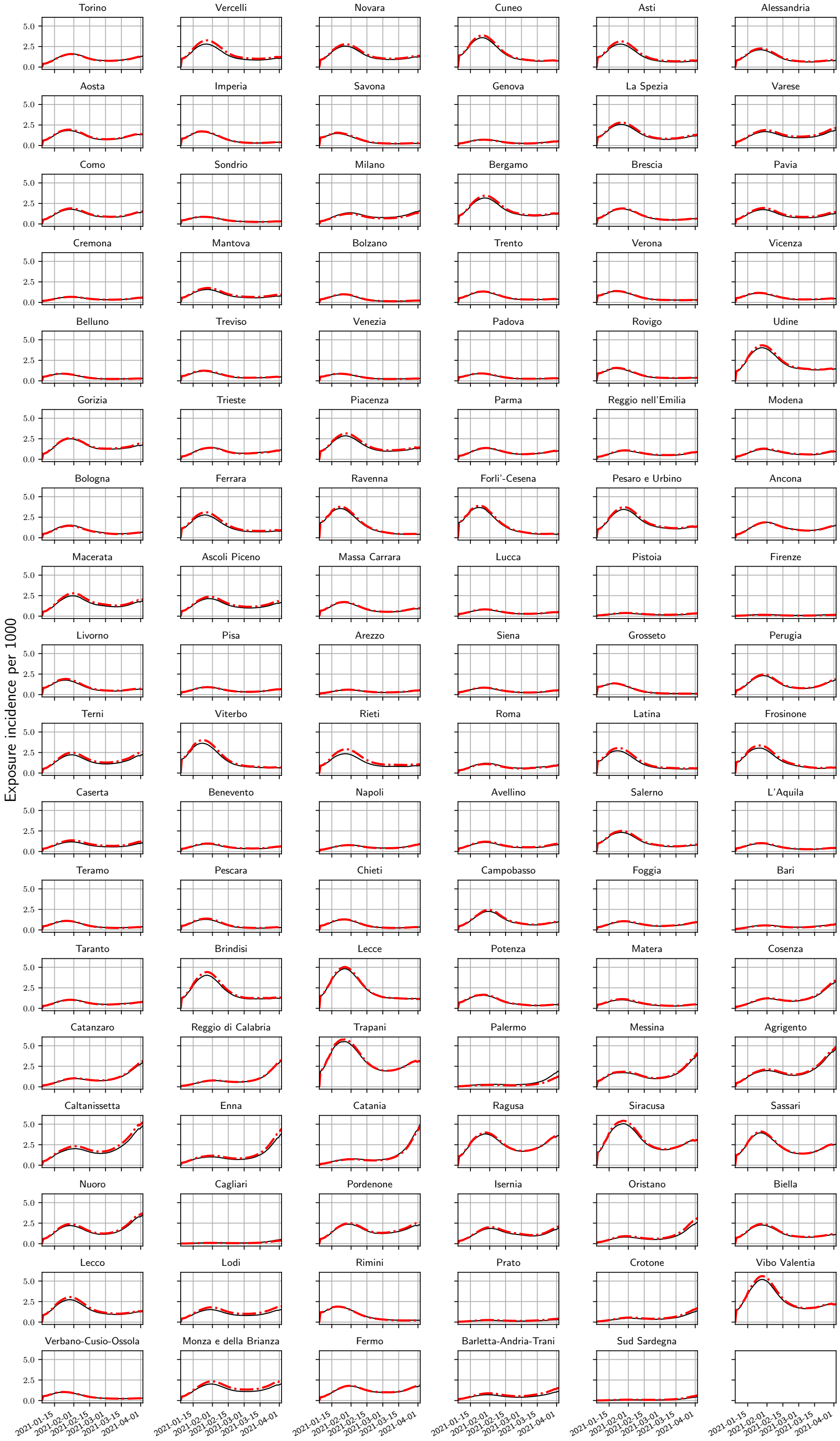


**Fig S3.** Comparison between the incidence in the exposed compartment $E$ (per 1'000 people) as evaluated by the model simplified for the optimal control (red) and the full epidemiological model (black). Results for the pessimistic scenario without vaccination. The difference is very small even for this very sensible compartment, justifying the simplifications undertaken.

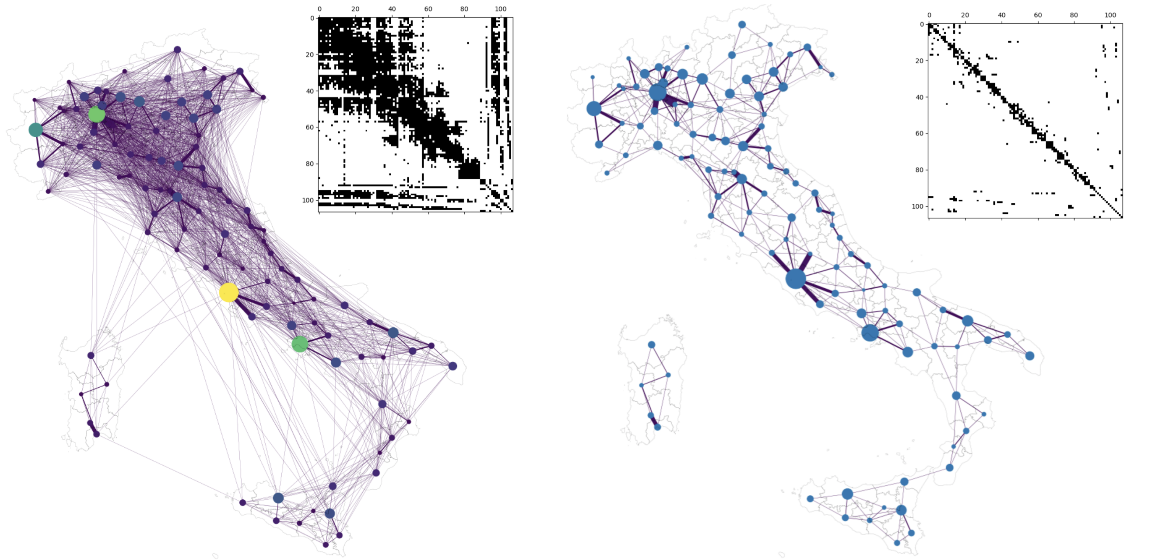

**Fig S4.** Simplification of the mobility matrix to obtain a sparse and tractable problem for the OCP. Note that, after computing the optimal vaccination strategy, we assess its effectiveness of on the full epidemiological model.

better track possible changes in the transmission rates, we adopt a data assimilation strategy based on an iterative particle filter [12] on a moving window to assess the values of the coefficients $\phi_i(t)$.

We initialize the model state variables and parameters using the results of a previous calibration effort [2] which fits the initial conditions, the baseline transmission $\beta_0$, as well as the coefficients $\epsilon_E$ and $\epsilon_I$ using a Bayesian framework for the period February 24 – May 1, 2020, on the basis of the official epidemiological bulletins released daily by Dipartimento della Protezione Civile [13] (data available online at `https://github.com/pcm-dpc/COVID-19`) and the bulletins of Epicentro, at Istituto Superiore di Sanità [14,15]. [2] presents three possible model calibrations based on three values of the symptomatic/asymptomatic infectious ratio ($\sigma = 0.5, 0.25, 0.10$). For our study we select the calibration results based on the central scenario, $\sigma = 0.25$.

In the data assimilation approach, the filter starts considering $N_r = 100$ model realizations at time $t_0$ (February 21st, 2020), whose state variables are $x_0^{(j)}, j = 1, \ldots, N_r$, where the superscript $(j)$ is the realization index and the subscript is the temporal index. Each realization is associated with a parameter combination that is randomly sampled from the posterior distribution evaluated in [2].

Note that all the epidemiological parameters estimated in [2], including the transmission rates, were spatially homogeneous, while possible temporal variations were imposed on fixed dates. Here instead temporal variations in the nodes' transmission rates are obtained by iteratively fitting coefficients $\phi_{k,i}^{(j)}$ against the regional hospitalization data on a moving window of $\tau = 14$ days. At time $t_0$, the coefficients $\phi_{0,i}^{(j)}$ are initialized by sampling from a truncated normal distribution (mean $\mu_0 = 1$, standard deviation 0.2, bounds 0.01-2). Knowing the state variables $x_k^{(j)}$ and the coefficients $\phi_{k,i}^{(j)}$ at time $t_k$, the latter having an ensemble mean $\mu_{k,i}$, we run the model for $\tau$ days by sampling new regional coefficients $\phi_{k+1,i}^{(j)}$ from the truncated normal distributions (mean $\mu_{k,i}$, standard deviation 0.2, bounds 0.01-2.0) assuming that the regional coefficients remain constant. The regional likelihood of each realization is then evaluated during the moving window assuming that the daily hospitalizations follow a gamma distribution (as in [2]). A resampling step (systematic resampling, see, e.g. [16]) selects and duplicates the coefficients $\tilde{\phi}_{k+1,i}^{(j)}$ associated with the largest likelihood values. These coefficients are then used to update the mean value $\mu_{k+1,i}$. This procedure is iterated four times on the same temporal window by sampling from a normal distribution with updated mean value $\mu_{k+1,i}$ and decreasing standard deviation up to 0.05. This set of coefficients is used to

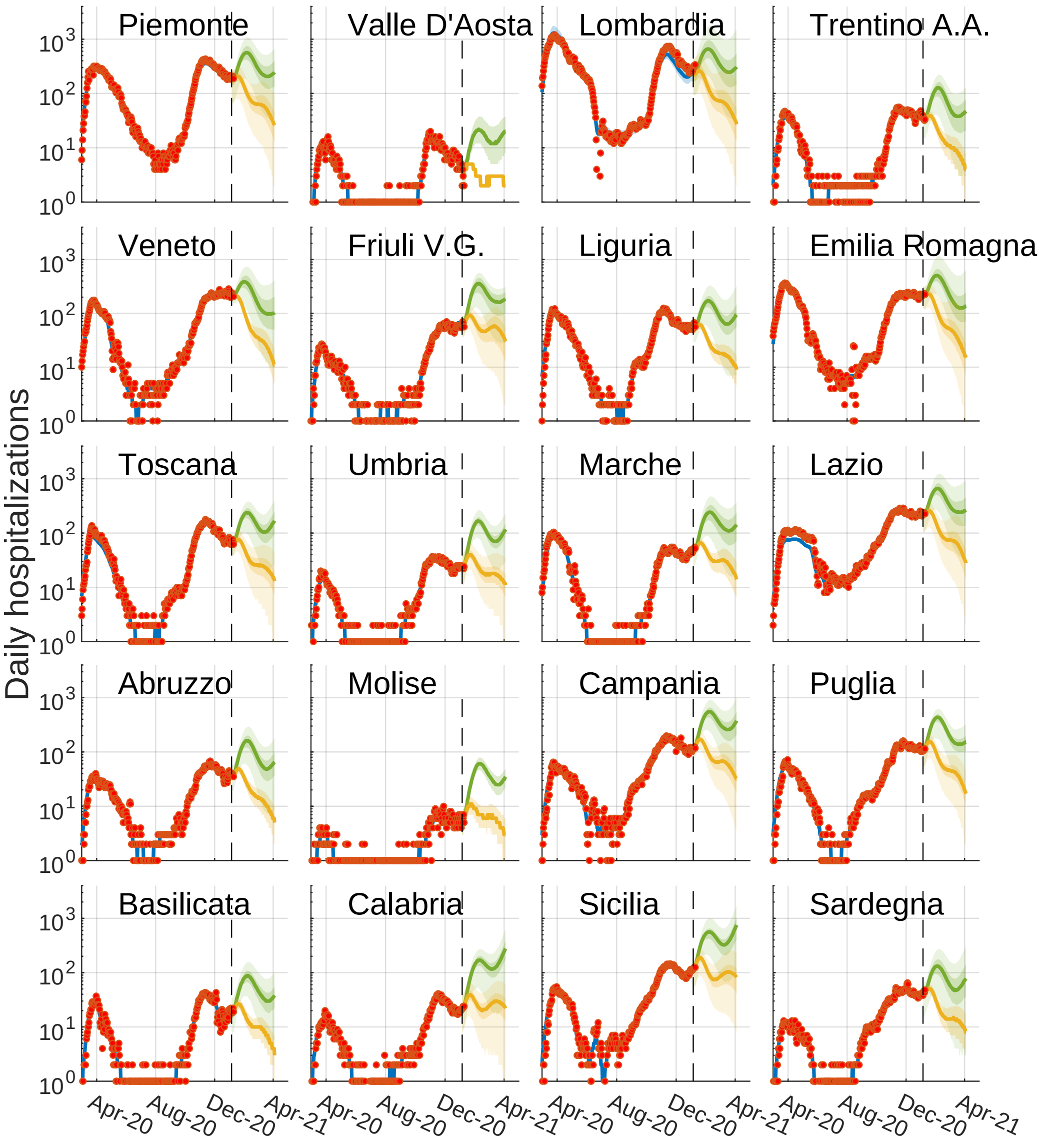


**Fig S5.** Modeled daily hospitalizations (blue) the versus hospitalization data (red dots), regional detail of Figure 2.A in the main text. The optimistic and pessimistic transmission scenarios are represented in green and yellow, respectively.

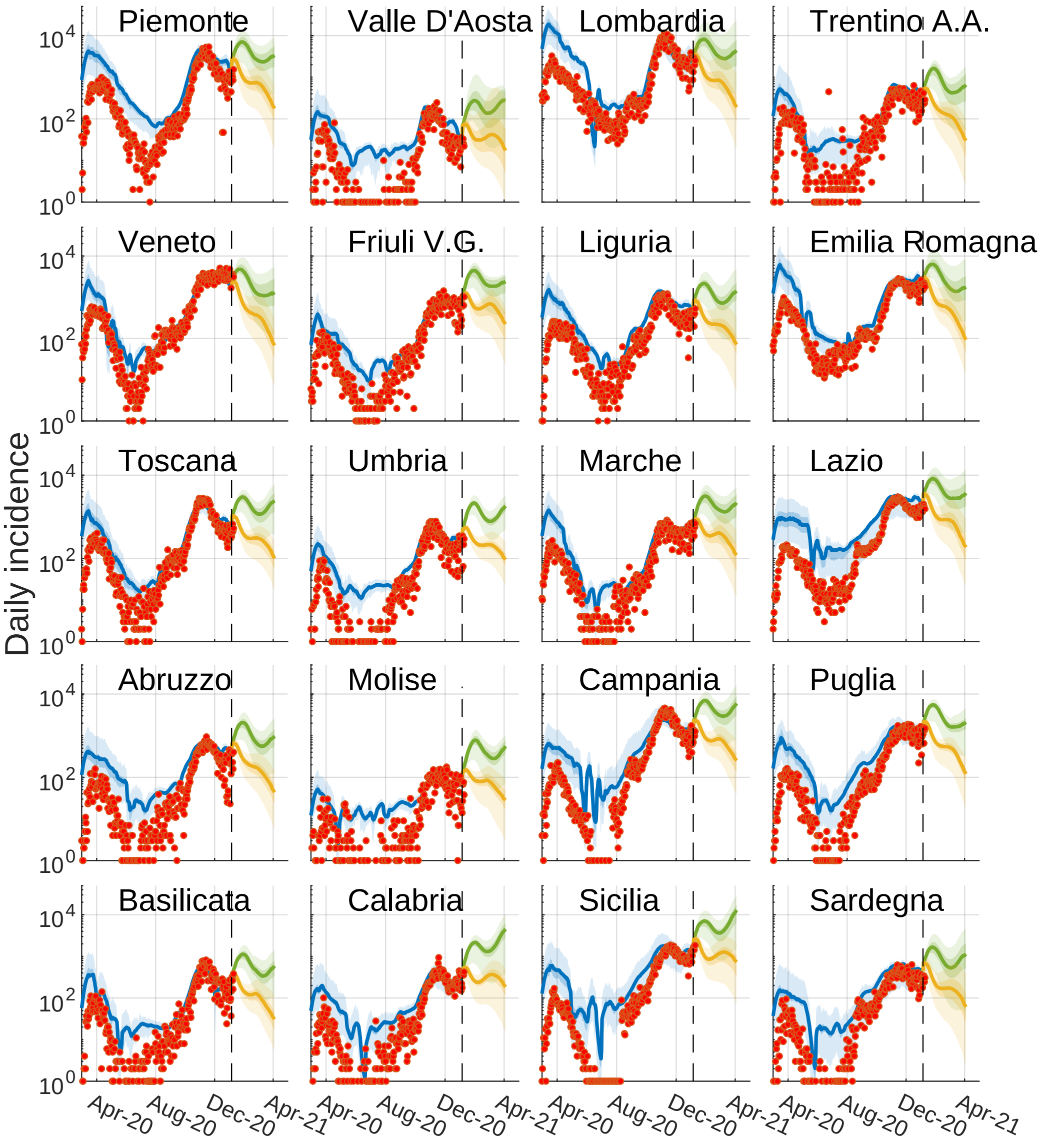


**Fig S6.** Modeled daily incidence (blue) versus the daily reported cases (red dots), regional detail of Figure 2.B in the main text. The optimistic and pessimistic transmission scenarios are represented in green and yellow, respectively.

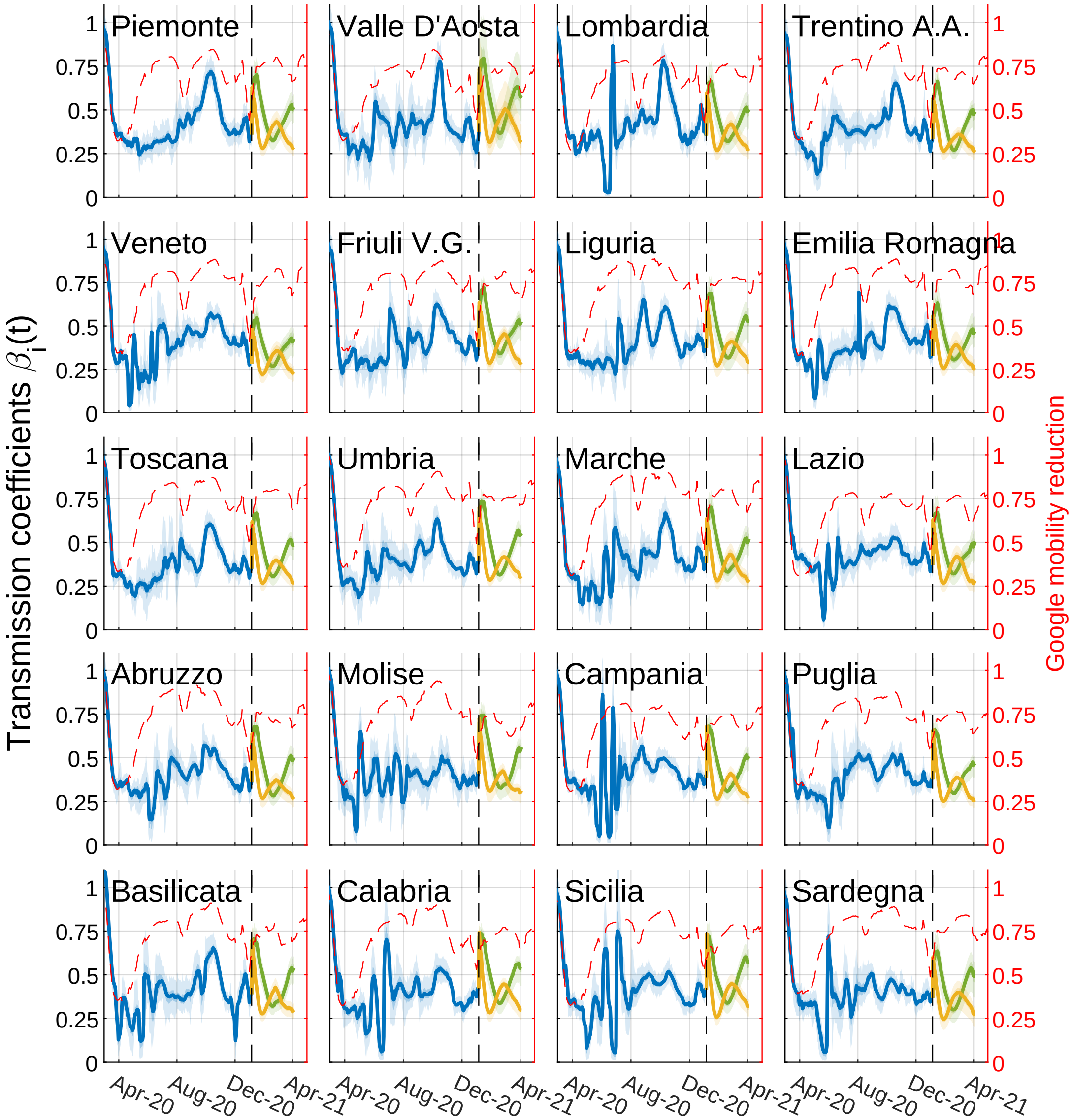


**Fig S7.** Values of the transmission parameters $\beta_i(t)$ (S6) as estimated in the data assimilation procedure (blue). The values used in the optimistic and pessimistic transmission scenarios are represented in green and yellow, respectively. The red lines represents the reduction in mobility and transmission achieved using google mobility data (coefficient $1 + g_i(t)/100$ in equation (S6)).

compute state variables and parameters at time $t_{k+1}$.

The assimilation is then repeat moving the window of one day, up to January 4, 2021 to produce the projections used in the main text. These projections are shown in Figure S8, which display the incidence for each province for the two scenarios. This view highlights the different trajectories between the optimistic and pessimistic scenarios and the spacial heterogeneities the optimal control framework optimizes on.

##### Spatial set-up

The modeling tools described in the following sections are applied to the Italian COVID-19 epidemic at the scale of second-level administrative divisions, i.e. provinces and metropolitan cities (currently, as of 2021, 107 spatial units). Official data about resident population at the provincial level is produced yearly by the Italian National Institute of Statistics (Istituto Nazionale di Statistica, ISTAT; data available at

`http://dati.istat.it/Index.aspx?QueryId=18460`). The latest update (January 1, 2019) has been used to inform the spatial distribution of the population.

The data to quantify nation-wide human mobility come from ISTAT (specifically, from the 2011 national census; data available online at `https://www.istat.it/it/archivio/139381`). Mobility fluxes, mostly reflecting commuting patterns related to work and study purposes, are provided at the scale of third-level administrative units (municipalities) [17,18]. These fluxes were upscaled to the provincial level following the administrative divisions of 2019, and used to evaluate the fraction $p_i$ of mobile people in each node $i$, as well as the fraction $q_{ij}$ of mobile people who move between $i$ and all other administrative units $j$ (see Supplementary Material in [1]).

#### Alternative strategies

We designed alternative vaccination strategies to compare the efficacy of the optimal solutions. Each strategy uses a decision variable, $\mathcal{V}_i$, as a basis for ranking the provinces and, thus, prioritize the allocation of vaccines. The decision variable is one of:

- **modelled future incidence**: the modelled total future incidence. This variable is updated daily during the projection reflecting the projected incidence knowing the effects of the already allocated vaccines;
- **modelled initial susceptibility**: the modelled number of susceptibles in each province at the start of the vaccination campaign;
- The aforementioned decision variables values are taken both absolute or relative to population e.g, one decision variable is **incidence** and another one is **incidence per habitant**, and the same for susceptibility.
- **the province's population**.
- **equal for all provinces**.

Once a decision variable is selected, there are two ways the doses might be distributed: focused and proportional.

- **Focused** Every day, we allocate $K = 1/7$th of the weekly stockpile delivery as follow: Every province is sorted (higher on top) according to its decision variable $\mathcal{V}_i$. We then allocate the maximum local rate $v_i^{max}$ to every province going down through the list, until the stockpile is empty. In other words, we find the province index $i$ that satisfy $\max_i \mathcal{V}_i$, and we assign to province $i$ $M_i = \min(v_i^{max}, K)$ vaccines. Then, we find the province $j$ that satisfy $\max_{j,j\neq i} \mathcal{V}_j$ and we assign it $M_j = \min(v_j^{max}, K - M_i)$. And so on, until no vaccine remains for allocation today. This strategy will concentrate the allocation on nodes with the highest values of the considered decision variable.

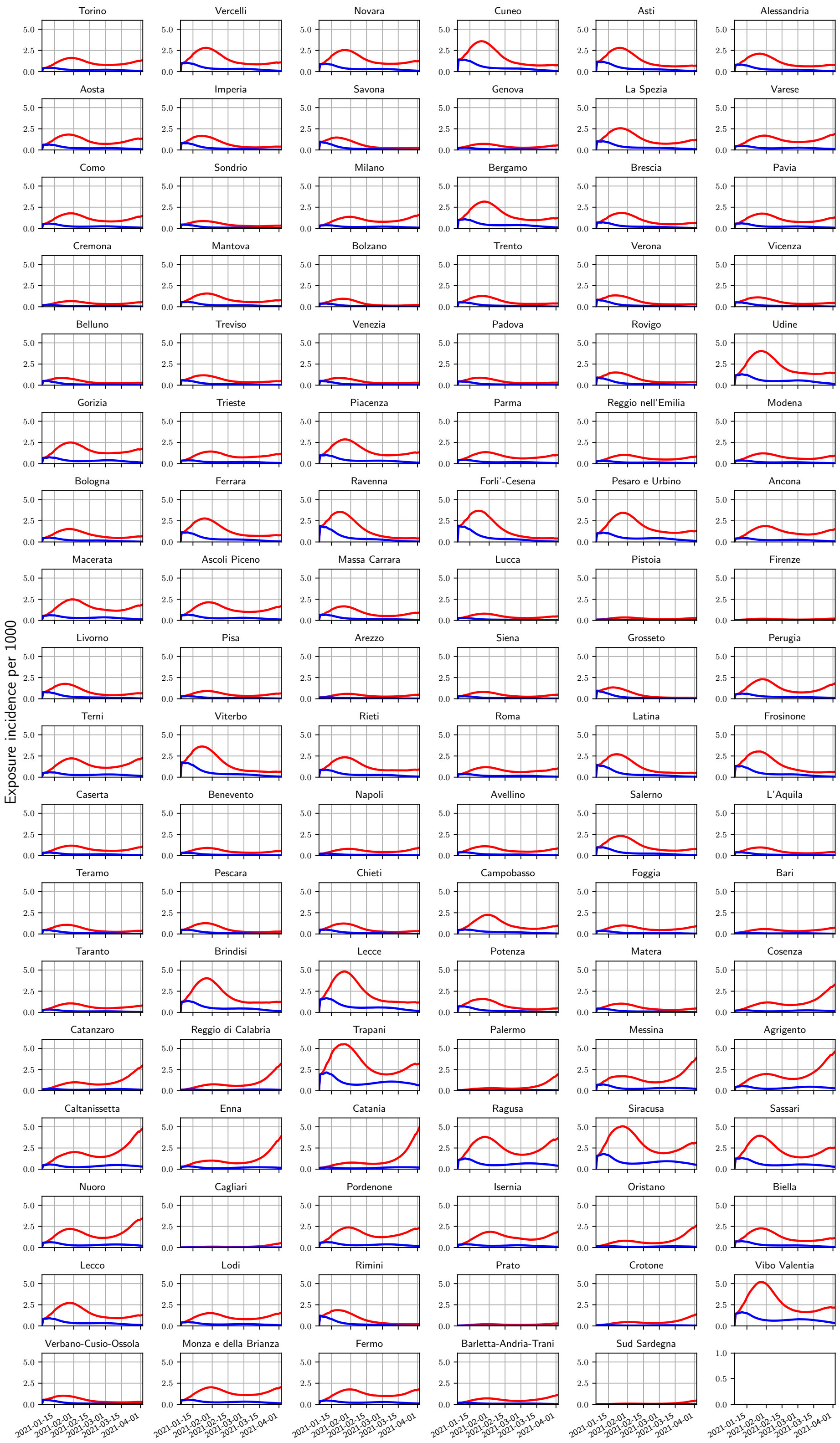


**Fig S8.** Projected incidence into the exposed compartment $E$ (per 1'000 people) for the pessimistic (red) and optimistic (blue) scenarios.

- **Proportional** In this case, assuming that on a given day there is a quantity of vaccine $K$ in the stockpile, we assign to each province $i$ an amount $M_i = \min(v_i^{max}, K \cdot \frac{\mathcal{V}_i}{\sum_j \mathcal{V}_j})$. This approach vaccinate each node proportionally to the value of its decision variable $\mathcal{V}_i$. Moreover, it exhibit very fast allocation as all vaccines are allocated the day following the stockpile delivery.

An additional strategy, proposed in [19] is developed in our benchmark. Named Greedy, it optimized the current projected impact of the allocation. Every week, after a delivery, we:

1. create 107 one week strategies, one per province. In each strategy one province receive as much as vaccine as possible (given susceptible remaining, stockpile and local logistic rate) for the seven days of the week.
2. Evaluate each of these strategies using the model, and compute the objective to minimize (number of infections).
3. Select the strategy that generate the highest reduction in the objective and keep it.
4. redo steps 1.–3. until no vaccines remains in the stockpile. Every iteration, keep the chosen strategies and re-apply it so it is taken into account.

The rational behind this strategy is that it optimize visible gains without requiring the full optimal control framework to be run.

### Additional results for the spatial model

We present the number of averted infections for each proposed vaccination strategies in Tables 1–2, and we show them side-by-side in Figure S9. The optimal solutions outperform all the others solutions. In fact, for every given posterior realisation, the optimal control solution always has the largest number of averted infections.

### Detailed analysis

To further investigate the features of the optimal solution, we present a linear scatter plot of the optimal proportion of vaccinated individuals per province (sorting variable) side by side with the province population, the projected incidence without vaccination, and the proportion of susceptible individuals at the start of the simulation. We present these results for the optimistic scenario in Figure S10 and for the pessimistic scenario in Figure S11. We find no clear visual pattern associating these covariates to the proportion of vaccinated individuals by the optimal strategy, highlighting again that the optimal allocation uses the epidemiological variables in a non-straightforward way, different from every alternative strategy we considered.

Finally, to highlight the temporal dimension of the prioritization strategy for the deployment of vaccine doses, we present a stackplot of the proportion of vaccine dose allocated in each province according to the optimal solution (Figure S12) and a heatmap (Figure S13) where the spatio-temporal pattern is compared for the different scenarios. From left to right, as we go to scenarios with higher deliveries, the optimal solution makes use of the new doses by both further re-enforcing already prioritized provinces (like a focused strategy) and by vaccinating new provinces (like a proportional strategy).

### Sensitivity Analysis

As seen in the Discussion, the optimal allocation is complex and highly specific, it uses every feature of the model to gain efficacy over other strategies. This raises the concern that the optimal

| Scenario | Vaccination strategy | Averted infections (Millions) | | Averted infections per dose | |
|---|---|---|---|---|---|
| | | Optimistic | Pessimistic | Optimistic | Pessimistic |
| 125'000 | Optimal | 0.146 | 0.672 | 0.0897 | 0.413 |
| | Incidence per hab. (focused) | 0.137 | 0.626 | 0.085 | 0.389 |
| | Incidence per hab. (proportional) | 0.106 | 0.509 | 0.0653 | 0.313 |
| | Incidence (proportional) | 0.103 | 0.494 | 0.0631 | 0.304 |
| | Greedy | 0.0951 | 0.388 | 0.0592 | 0.242 |
| | Equal (proportional) | 0.0749 | 0.395 | 0.0461 | 0.243 |
| | Susceptible per hab. (proportional) | 0.074 | 0.393 | 0.0456 | 0.242 |
| | Population (proportional) | 0.0691 | 0.387 | 0.0425 | 0.238 |
| | Susceptible (proportional) | 0.068 | 0.384 | 0.0419 | 0.236 |
| | Incidence (focused) | 0.0596 | 0.315 | 0.0371 | 0.196 |
| | Susceptible per hab. (focused) | 0.0328 | 0.213 | 0.0204 | 0.133 |
| | Susceptible (focused) | 0.0299 | 0.185 | 0.0186 | 0.115 |
| | Population (focused) | 0.0288 | 0.176 | 0.0179 | 0.109 |
| | Equal (focused) | 0.0259 | 0.157 | 0.0161 | 0.0977 |
| 250'000 | Optimal | 0.228 | 1.1 | 0.0701 | 0.34 |
| | Incidence per hab. (focused) | 0.214 | 1.03 | 0.0666 | 0.321 |
| | Incidence per hab. (proportional) | 0.18 | 0.893 | 0.0554 | 0.275 |
| | Incidence (proportional) | 0.174 | 0.896 | 0.0535 | 0.276 |
| | Equal (proportional) | 0.141 | 0.739 | 0.0433 | 0.227 |
| | Susceptible per hab. (proportional) | 0.139 | 0.734 | 0.0428 | 0.226 |
| | Population (proportional) | 0.132 | 0.735 | 0.0407 | 0.226 |
| | Susceptible (proportional) | 0.13 | 0.729 | 0.0401 | 0.224 |
| | Greedy | 0.117 | 0.653 | 0.0365 | 0.203 |
| | Incidence (focused) | 0.116 | 0.581 | 0.0359 | 0.181 |
| | Susceptible per hab. (focused) | 0.0719 | 0.449 | 0.0224 | 0.14 |
| | Equal (focused) | 0.0708 | 0.369 | 0.022 | 0.115 |
| | Susceptible (focused) | 0.053 | 0.335 | 0.0165 | 0.104 |
| | Population (focused) | 0.0393 | 0.232 | 0.0122 | 0.0723 |
| 479'700 | Optimal | 0.334 | 1.7 | 0.0535 | 0.272 |
| | Incidence per hab. (focused) | 0.318 | 1.6 | 0.0515 | 0.259 |
| | Incidence per hab. (proportional) | 0.282 | 1.45 | 0.0452 | 0.232 |
| | Incidence (proportional) | 0.274 | 1.44 | 0.044 | 0.231 |
| | Equal (proportional) | 0.241 | 1.28 | 0.0387 | 0.205 |
| | Susceptible per hab. (proportional) | 0.24 | 1.28 | 0.0384 | 0.205 |
| | Population (proportional) | 0.232 | 1.29 | 0.0373 | 0.206 |
| | Susceptible (proportional) | 0.229 | 1.28 | 0.0368 | 0.205 |
| | Incidence (focused) | 0.203 | 1.05 | 0.0329 | 0.17 |
| | Greedy | 0.203 | 1.14 | 0.0329 | 0.186 |
| | Susceptible per hab. (focused) | 0.138 | 0.869 | 0.0225 | 0.141 |
| | Equal (focused) | 0.126 | 0.635 | 0.0204 | 0.103 |
| | Susceptible (focused) | 0.0885 | 0.56 | 0.0143 | 0.0907 |
| | Population (focused) | 0.0863 | 0.536 | 0.014 | 0.0869 |

**Table 1. Absolute number of averted infections** for the scenarios with the lower weekly stockpile delivery.

allocation might be unstable: if there are errors in model projections, it might perform significantly worth than other (less specific) strategies.

We perform a sensitivity analysis of the optimal allocation. Instead of evaluating the performance on other posterior realizations like in Figure S9, we evaluate it on completely different projections. These 100 projections are obtained by shuffling each provinces projected dynamics (shown in Figure S8). The results are shown in Figure S14. As expected, we observed degraded performance for all strategies, but the optimal control strategy does show a particular fragility under these conditions.

| Scenario | Vaccination strategy | Averted infections (Millions) | | Averted infections per dose | |
|---|---|---|---|---|---|
| | | Optimistic | Pessimistic | Optimistic | Pessimistic |
| 1M | Optimal | 0.484 | 2.54 | 0.0372 | 0.196 |
| | Incidence per hab. (focused) | 0.467 | 2.44 | 0.0363 | 0.19 |
| | Incidence per hab. (proportional) | 0.437 | 2.31 | 0.0336 | 0.177 |
| | Incidence (proportional) | 0.431 | 2.3 | 0.0332 | 0.177 |
| | Equal (proportional) | 0.402 | 2.15 | 0.031 | 0.165 |
| | Susceptible per hab. (proportional) | 0.401 | 2.15 | 0.0309 | 0.165 |
| | Population (proportional) | 0.399 | 2.18 | 0.0307 | 0.168 |
| | Susceptible (proportional) | 0.397 | 2.18 | 0.0306 | 0.167 |
| | Incidence (focused) | 0.369 | 1.94 | 0.0287 | 0.151 |
| | Greedy | 0.354 | 1.88 | 0.0275 | 0.146 |
| | Susceptible per hab. (focused) | 0.287 | 1.59 | 0.0223 | 0.123 |
| | Equal (focused) | 0.212 | 1.12 | 0.0165 | 0.0872 |
| | Susceptible (focused) | 0.171 | 1.06 | 0.0133 | 0.0823 |
| | Population (focused) | 0.166 | 0.996 | 0.0129 | 0.0775 |
| 1.5M | Optimal | 0.572 | 3.03 | 0.0293 | 0.156 |
| | Incidence per hab. (focused) | 0.557 | 2.94 | 0.0289 | 0.153 |
| | Incidence per hab. (proportional) | 0.536 | 2.85 | 0.0275 | 0.146 |
| | Incidence (proportional) | 0.531 | 2.84 | 0.0272 | 0.146 |
| | Population (proportional) | 0.507 | 2.75 | 0.026 | 0.141 |
| | Equal (proportional) | 0.507 | 2.71 | 0.026 | 0.139 |
| | Susceptible per hab. (proportional) | 0.507 | 2.71 | 0.026 | 0.139 |
| | Susceptible (proportional) | 0.506 | 2.74 | 0.026 | 0.141 |
| | Incidence (focused) | 0.492 | 2.6 | 0.0255 | 0.135 |
| | Greedy | 0.451 | 2.38 | 0.0234 | 0.124 |
| | Susceptible per hab. (focused) | 0.402 | 2.15 | 0.0209 | 0.112 |
| | Equal (focused) | 0.296 | 1.62 | 0.0154 | 0.084 |
| | Susceptible (focused) | 0.269 | 1.58 | 0.014 | 0.0821 |
| | Population (focused) | 0.245 | 1.46 | 0.0127 | 0.0755 |
| 2M | Optimal | 0.632 | 3.35 | 0.0243 | 0.129 |
| | Incidence per hab. (focused) | 0.62 | 3.29 | 0.0241 | 0.128 |
| | Incidence per hab. (proportional) | 0.606 | 3.22 | 0.0232 | 0.123 |
| | Incidence (proportional) | 0.602 | 3.21 | 0.0231 | 0.123 |
| | Population (proportional) | 0.583 | 3.14 | 0.0224 | 0.121 |
| | Susceptible per hab. (proportional) | 0.583 | 3.12 | 0.0223 | 0.119 |
| | Susceptible (proportional) | 0.583 | 3.14 | 0.0224 | 0.121 |
| | Equal (proportional) | 0.582 | 3.11 | 0.0222 | 0.118 |
| | Incidence (focused) | 0.58 | 3.07 | 0.0225 | 0.12 |
| | Greedy | 0.538 | 2.77 | 0.0209 | 0.108 |
| | Susceptible per hab. (focused) | 0.493 | 2.63 | 0.0192 | 0.102 |
| | Equal (focused) | 0.438 | 2.36 | 0.017 | 0.0919 |
| | Susceptible (focused) | 0.366 | 2.11 | 0.0142 | 0.0819 |
| | Population (focused) | 0.35 | 2.02 | 0.0136 | 0.0785 |

**Table 2. Absolute number of averted infections** for the scenarios with the largest weekly stockpile deliveries.

2. Bertuzzo E, Mari L, Pasetto D, Miccoli S, Casagrandi R, Gatto M, et al. The geography of COVID-19 spread in Italy and implications for the relaxation of confinement measures.

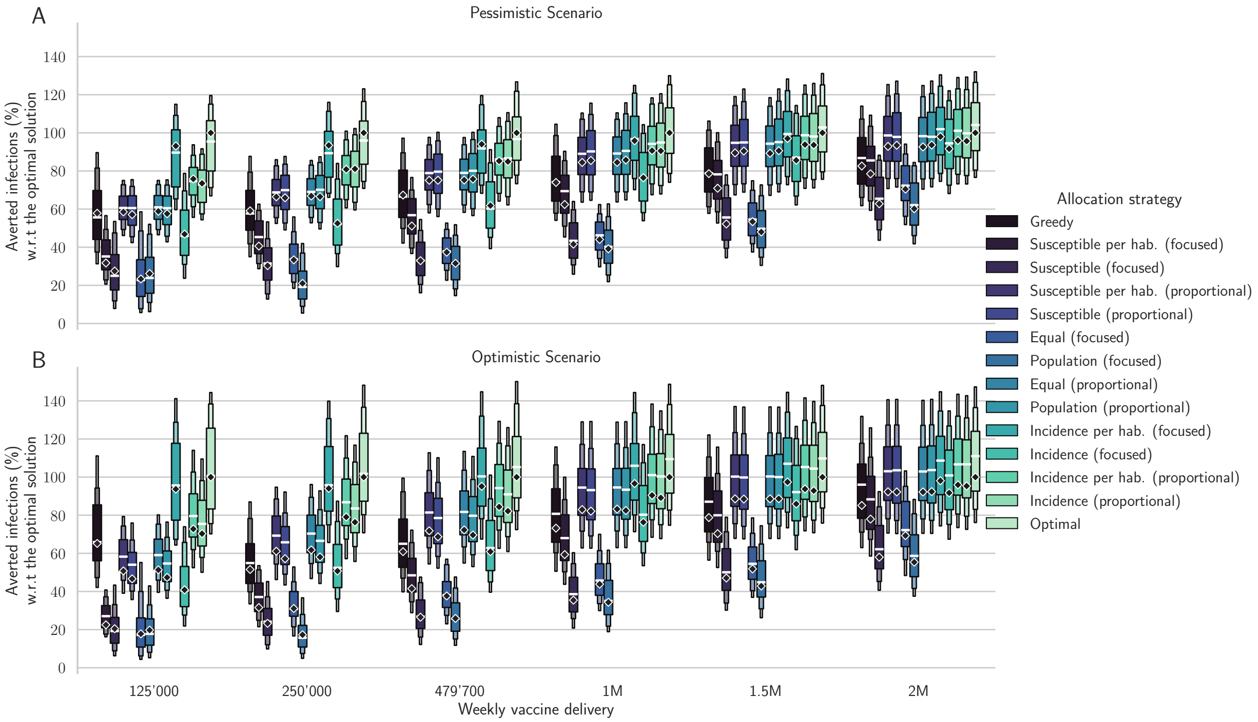


**Fig S9.** **Comparison of different allocation strategies**. Percentages of averted infections per vaccine dose from January 4, 2021 to April 4, 2021 using different vaccine distribution strategies for the pessimistic (panel A) and the optimistic (panel B) scenario based on: the optimal solution, the spatial distribution of the population, the amount of susceptible individuals at the beginning of the vaccination campaign, and the projected disease incidence in the absence of control. We optimize a median realization of the modeled posterior (diamonds), and assess the performance on the whole posterior (box plots). The results are normalized by the number of averted infections in the optimized solution (see Tables 1–2 for absolute values).


Nature Communications. 2020;11(1):4264. doi:10.1038/s41467-020-18050-2.

8. Rawlings JB, Mayne DQ, Diehl M. Model Predictive Control: Theory, Computation, and Design. 2nd ed. Nob Hill Publishing; 2017.

9. Andersson J, Åkesson J, Diehl M. CasADi: A Symbolic Package for Automatic Differentiation and Optimal Control. In: Recent Advances in Algorithmic Differentiation. Lecture Notes in

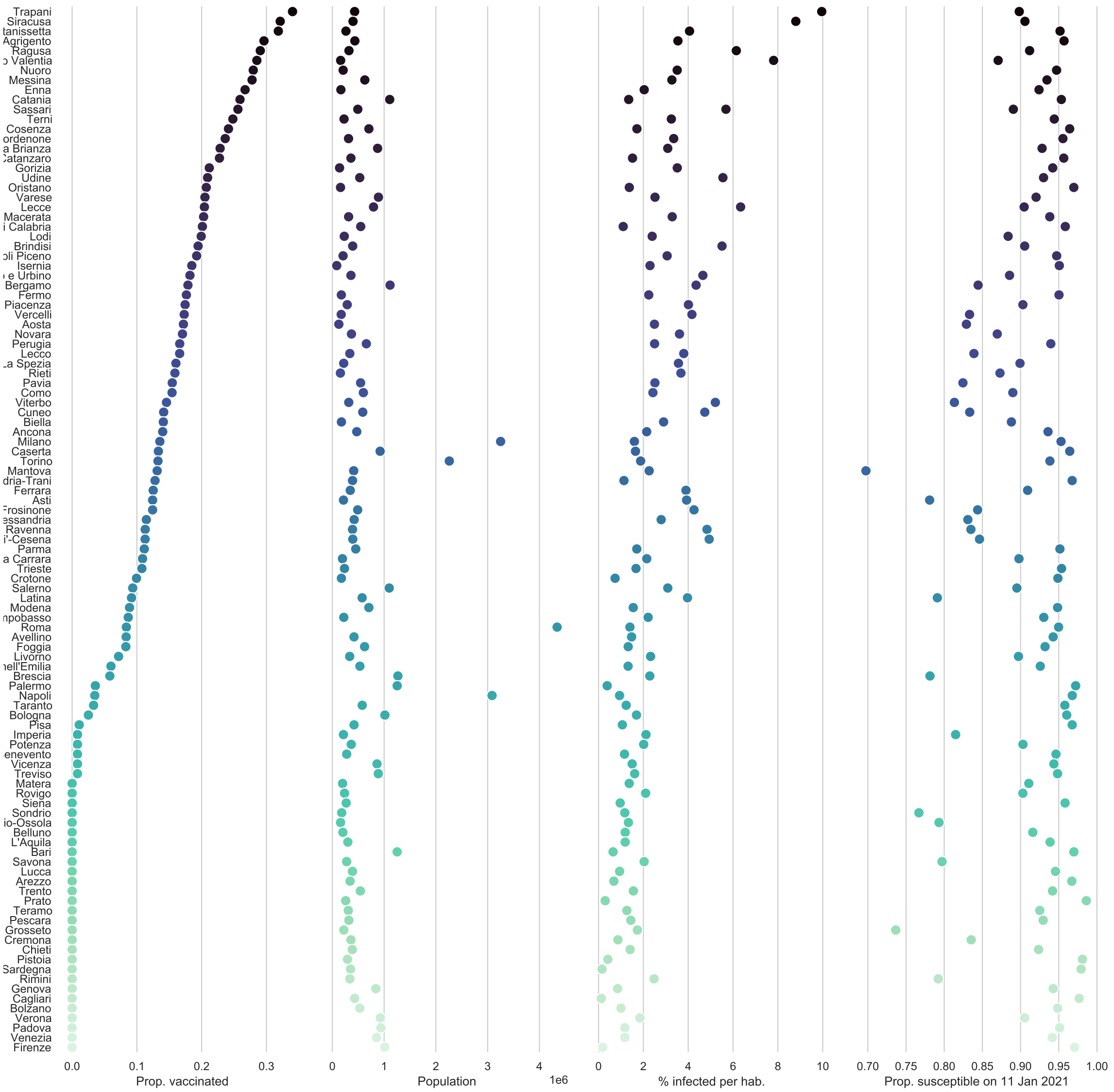


**Fig S10. Control and covariates for the optimistic scenario with a stockpile delivery of 479’700 vaccine doses.**


Computational Science and Engineering. Springer, Berlin, Heidelberg; 2012. p. 297–307. Available from: `https://link.springer.com/chapter/10.1007/978-3-642-30023-3_27`.

10. Wächter A, Biegler LT. On the implementation of an interior-point filter line-search algorithm for large-scale nonlinear programming. Mathematical Programming. 2006;106(1):25–57. doi:10.1007/s10107-004-0559-y.

11. HSL. A collection of Fortran codes for large scale scientific computation.;. Available from: `http://www.hsl.rl.ac.uk/`.

12. Manoli G, Rossi M, Pasetto D, Deiana R, Ferraris S, Cassiani G, et al. An iterative particle filter approach for coupled hydro-geophysical inversion of a controlled infiltration experiment. Journal of Computational Physics. 2015;283(C):37–51. doi:10.1016/j.jcp.2014.11.035.

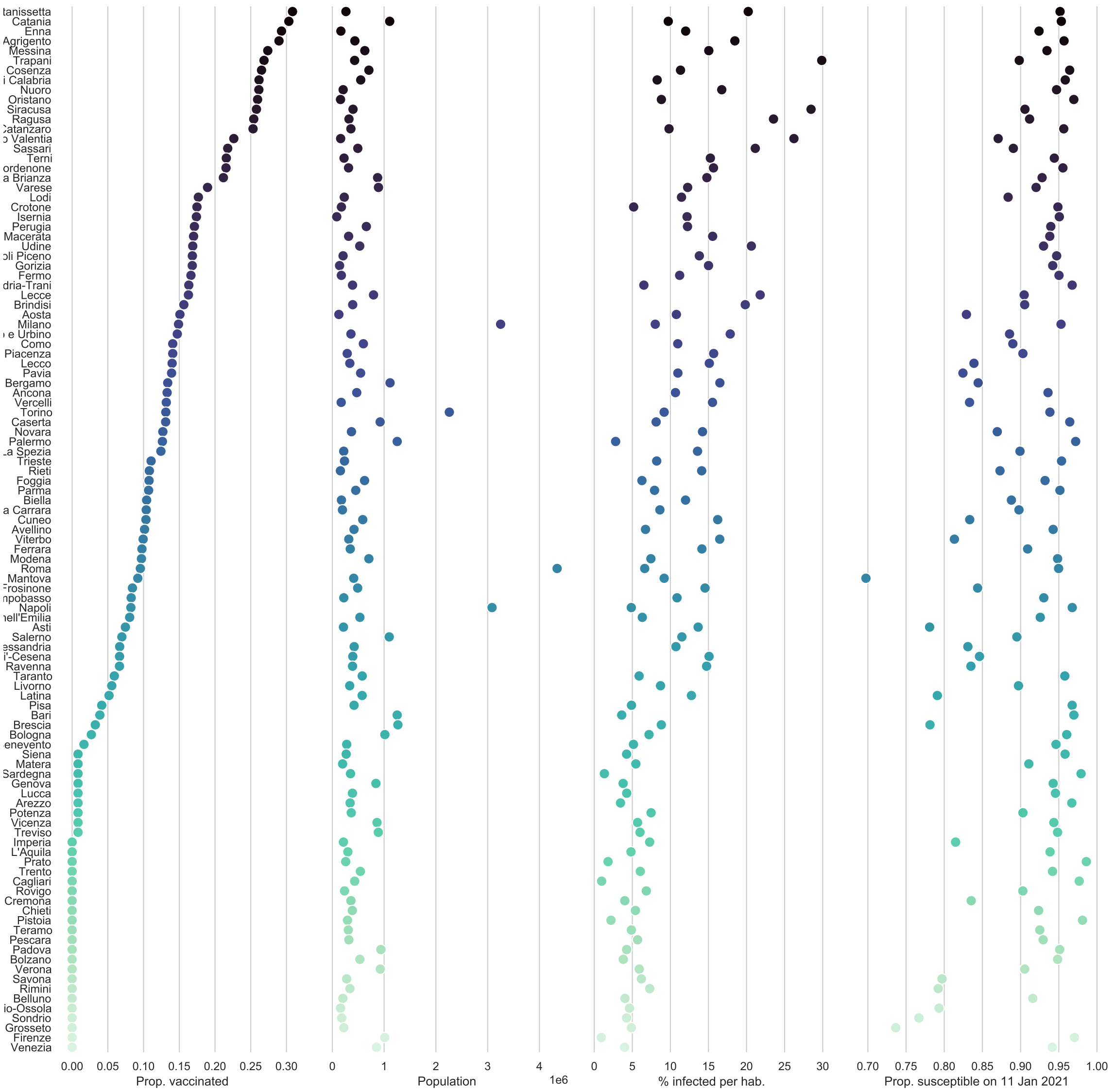


**Fig S11.** **Control and covariates for the pessimistic scenario with a stockpile delivery of 479'700 vaccine doses.**


13. Dipartimento della Protezione Civile. Emergenza Coronavirus: la risposta nazionale;. Available from: `http://www.protezionecivile.gov.it/attivita-rischi/rischio-sanitario/emergenze/coronavirus`.

14. Istituto Superiore di Sanità. Coronavirus: ultimi aggiornamenti; 2020. Available from: `https://www.epicentro.iss.it/coronavirus/aggiornamenti`.

15. Palmieri L, Andrianou X, Barbariol P, Bella A, Bellino S, Benelli E, et al.. Characteristics of COVID-19 patients dying in Italy Report based on available data on October 4th, 2020; 2020. Available from: `https://www.epicentro.iss.it/en/coronavirus/sars-cov-2-analysis-of-deaths`.

16. Douc R, Cappe O. Comparison of resampling schemes for particle filtering. In: ISPA 2005. Proceedings of the 4th international symposium on image and signal processing and analysis,

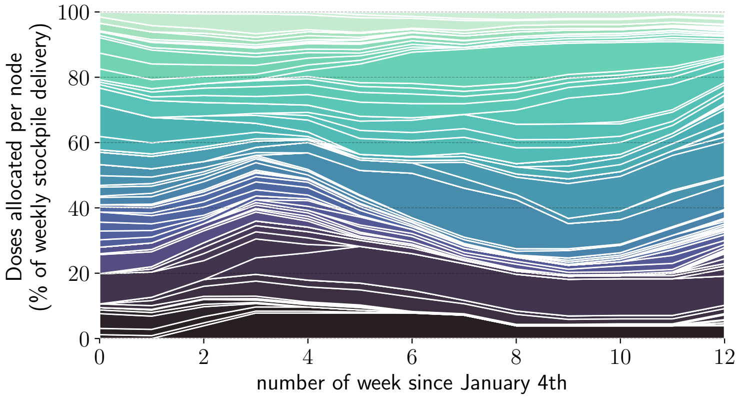


**Fig S12.** **Time allocation** for the pessimistic scenario with a stockpile delivery of 479'700. We see for each week, how the 479'700 doses are spread across the provinces, as percent. This view unravel the temporal pattern in the allocation.

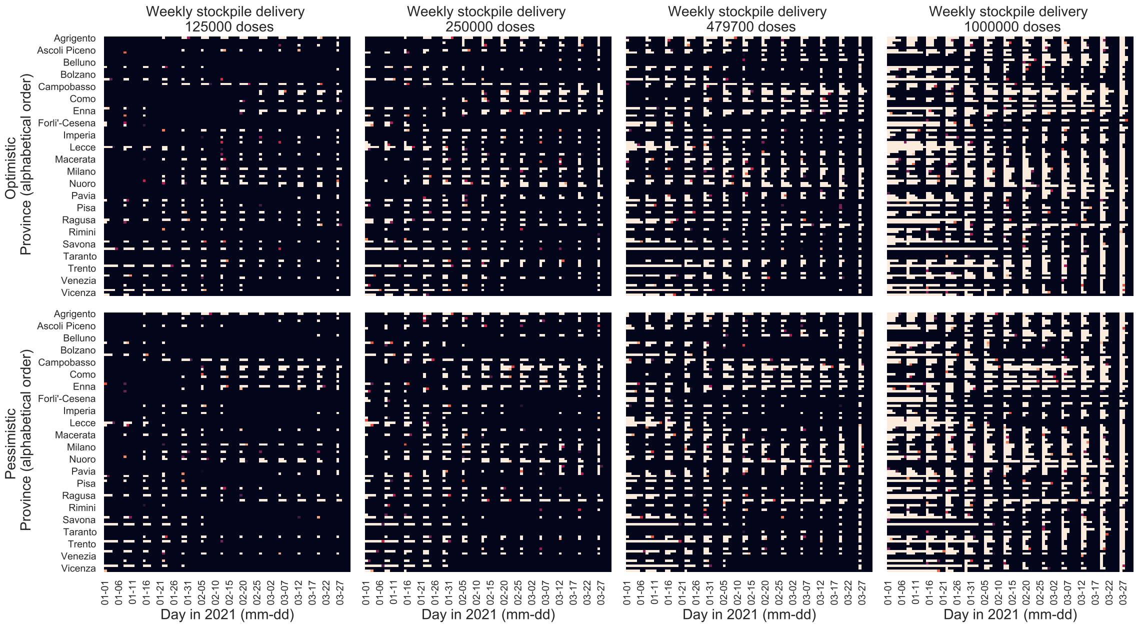


**Fig S13.** **Heatmap** showing the allocation in space and time for different weekly delivery scenarios (left to right) and different transmission scenario (Optimistic at the top, Pessimistic at the bottom). The x-axis represent time (one square per day) and the y-axis space (one square per province), and the color represents the proportion of individuals vaccinated on this day in this province by the optimal solution, with black meaning no vaccination while yellow display the maximum logistic rate in this province per inhabitant (which is equal for all provinces).


2005.; 2005. p. 64–69.

17. Pepe E, Bajardi P, Gauvin L, Privitera F, Lake B, Cattuto C, et al. COVID-19 outbreak response, a dataset to assess mobility changes in Italy following national lockdown. Scientific Data. 2020;7(1). doi:10.1038/s41597-020-00575-2.

18. Vollmer MAS, others. Report 20: Using mobility to estimate the transmission intensity of COVID-19 in Italy: A subnational analysis with future scenarios. Imperial College London; 2020. Available from: https://www.imperial.ac.uk/mrc-global-infectious-disease-analysis/covid-19/report-20-italy/.

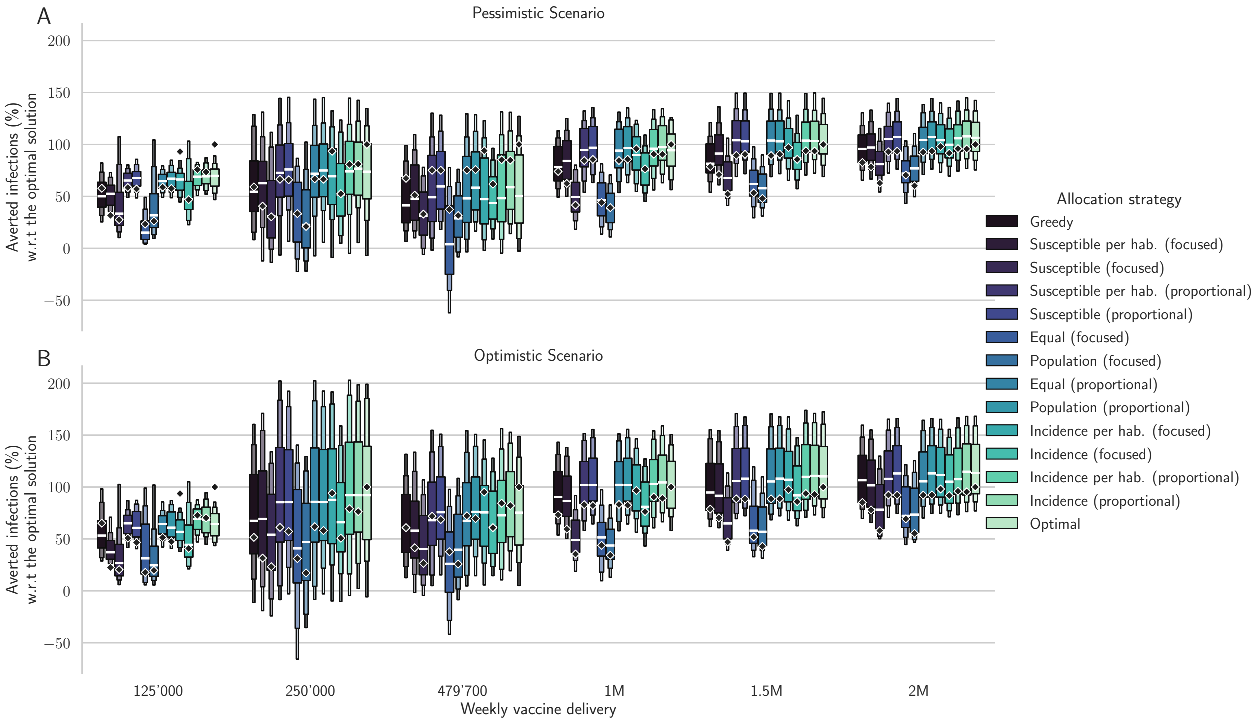


**Fig S14. Sensitivity analysis of different allocation strategies**. Percentages of averted infections per vaccine dose from January 4, 2021 to April 4, 2021 using different vaccine distribution strategies for the pessimistic (panel A) and the optimistic (panel B) scenario for all alternative strategies. Here the median realization of the modeled posterior is optimized (diamonds), and the comparison is done on shuffled dynamics (random allocation of each province dynamics to another province, box plots). The results are normalized by the number of averted infections in the optimized solution (see Table 1-2 for absolute values).

19. Venkatramanan S, Chen J, Fadikar A, Gupta S, Higdon D, Lewis B, et al. Optimizing spatial allocation of seasonal influenza vaccine under temporal constraints. PLOS Computational Biology. 2019;15(9):e1007111. doi:10.1371/journal.pcbi.1007111.
